# Health and socioeconomic characteristics underlying educational attainment of children born preterm in England: a population cohort study using linked data

**DOI:** 10.1101/2025.11.13.25340148

**Authors:** Sadia Haider, Athanasios Tsanas, G. David Batty, Rebecca M Reynolds, Melvyn Roffe, Heather C Whalley, Riccardo E Marioni, Hilary Richardson, Cheryl Battersby, James P Boardman

## Abstract

**Background:** Preterm birth (PTB) is a leading cause of atypical brain development and cognitive impairment, however, there are sparse data on its impact on statutory educational assessments. We aimed to describe school readiness at 5 years and educational attainment at 6–7 years of children born preterm in England and identify the early life factors that associate with attainment.

**Methods:** We created a novel record linkage between the National Neonatal Research Database and the National Pupil Database for all babies born <32 weeks’ gestation in England (2008–2012) who survived to neonatal discharge. We used logistic regression to investigate associations between clinical and demographic variables and performance in six statutory school-age assessments.

**Findings:** Data from 15,857 children were analysed (53.3% boys). Fifty-seven percent did not meet the school readiness level at age 5, 51% did not meet expected attainment at ages six to seven in writing, maths (48%), reading (42%), and science (36%). Children born at 23–24 weeks had two to three times higher odds of not meeting expected levels compared to those born at 31 weeks (age 5: adjusted odds ratio 2.86, 95% CI 2.19-3.73). Children born in the most deprived areas at birth had 1 7 to 2-fold the risk of under-attainment compared to those in the least deprived. In adjusted models, male sex and season of birth conferred increased risk, alongside several potentially mutable risks: smoking during pregnancy, exposure to ante-or postnatal corticosteroids, severe acquired neonatal brain injuries, co-morbidities of PTB, nutrition during neonatal care, and childhood deprivation.

**Interpretation:** PTB poses a substantial risk for educational under-attainment at ages 5 through 7 years, particularly when combined with socioeconomic deprivation. Addressing neighbourhood-and family-level social inequalities should be given similar priority as reducing medical complications of PTB for improving the educational attainment of children born preterm.

**Funding:** UKRI Medical Research Council

**RESEARCH IN CONTEXT:** *Evidence before this study:* We searched PubMed from October 1^st^, 2015, to October 1^st^, 2025, with no language restrictions for publications using the terms ((“preterm” OR “premature” OR “gestational age”) AND “socioeconomic” AND (“school” OR “education”)) in the title/abstract. Our search identified 20 relevant peer-reviewed studies, including one narrative systematic review that examined how socioeconomic inequalities and preterm birth interact to modify health and education outcomes. Three studies were conducted in the UK and used linked data to investigate educational outcomes for preterm-born children. The remainder were conducted in Scandinavian countries, Canada, Australia, and the Netherlands. Collectively, these studies have consistently shown that children born preterm have lower educational attainment compared with those born at term: a well-established dose-response relationship exists between gestational age and educational outcomes, with lower gestational age associated with progressively poorer attainment across all stages of education. However, most existing linkage studies did not capture the clinical heterogeneity of the preterm-born population underlying this gradient, as these data are rarely available in population-level datasets linked to education records. Whilst some studies included measures of socioeconomic status as a covariate, their independent effects have seldom been investigated, particularly relative to neonatal risk factors.

*Added value of this study:* The present study addresses these gaps through a novel linkage of population-level neonatal clinical data with national statutory educational assessments. We describe the high prevalence of low attainment among very preterm children (<32 weeks’ gestation) at ages 5 through 7 years in England. The linkage enables a granular characterisation of how maternal and neonatal conditions, exposures and treatments, and neighbourhood-and family-level measures of social disadvantage explain educational performance of children born <32 weeks’ gestation. The findings show a dose effect of low GA on low attainment and that the adverse effects of socioeconomic deprivation at birth persist through early school age. The effect of family-level deprivation at the start of school age is comparable in magnitude to that of severe neonatal brain injury, however, impacts a greater proportion of the population. As such, this study provides a more holistic understanding of the pathways underlying educational risk in this population.

*Implications of all the available evidence:* Improving the educational attainment of very preterm children is likely to require a reduction in social inequalities in pregnancy and early childhood, alongside care practices and research to minimise comorbidities of preterm birth. Furthermore, the identification of modifiable exposures, including maternal non-smoking, antenatal or postnatal corticosteroid exposure, breastfeeding, and deferred school entry or targeted additional support for very preterm children born in the summer months, offers practical targets for parents, clinicians, teachers and school leaders and educators, that could facilitate long-term educational attainment for children born preterm.

## Introduction

Globally, preterm birth (PTB) affects 13 4 million pregnancies per annum^1^. Over the past two decades, the survival rate of children born preterm has improved, but 10-15% of children born very preterm (<32 weeks) develop cerebral palsy, and 30-50% develop an intellectual or behavioural disability.^2^ Furthermore, preterm birth is more common among socioeconomically disadvantaged groups, reflecting social gradients in maternal health, environmental exposures, and access to antenatal care.^3^ Consequently, preterm children face a double burden of biological immaturity and social disadvantage, with the latter influencing developmental outcomes through limited access to resources, greater exposure to stress, and reduced opportunities for cognitive stimulation. There is a paucity of data on the relationship between preterm birth and statutory educational assessments, and on the relative importance of socioeconomic disadvantage in shaping this relationship, particularly around the transition to school when cognitive demands substantially increase. Statutory assessments often determine the level of support provided to children.

At the individual level, some very preterm children are free of neurodevelopmental and cognitive impairments, indicating that it is not low gestational age (GA) alone that confers risk. We hypothesise that risks which commonly co-occur with PTB influence the susceptibility to injury and dysmaturation and may explain differences in attainment at school age. These preterm birth-associated risk factors (PTB-RFs) are biological, psychosocial, and social/infrastructural. They can affect parent or child, or be shared, for instance: alterations in the perinatal stress environment,^4,5^ systemic inflammation,^6,7^ suboptimal neonatal nutrition,^8^ complications of PTB, such as bronchopulmonary dysplasia and necrotising enterocolitis,^9^ and social inequalities.^3,10,11^ Understanding the relative contributions of PTB-RFs to important educational outcomes is necessary to inform clinical interventions and social policies designed to facilitate the educational attainment of children born preterm.

The National Neonatal Research Database (NNRD) contains data on all neonatal admissions across the United Kingdom (UK) from 2007 to the present.^12^ The National Pupil Database (NPD) is an administrative dataset of educational outcomes collected and maintained by the Department for Education (DfE) in England.^13^ We created a novel record linkage between the NNRD and the NPD^14^ to quantify the proportion of preterm children who do not meet expected levels in statutory educational assessments and test the hypothesis that specific PTB-RFs modify the relationship between low GA and outcomes for school readiness and educational attainment at 5 to 7 years among children growing up in England.

## Methods

### Data source

The study links data from the National Neonatal Research Database (NNRD)^10^ and National Pupil Database (NPD)^11,14^. See Supplement for details of the datasets.

### Study population

This retrospective cohort comprised children born and cared for in a neonatal unit (NNU) in England between 1st September 2008 and 31st August 2012, with a recorded GA below 32 completed weeks, who survived to discharge, and had a linked neonatal and education record. The study epoch was chosen to include children old enough to be eligible for assessments at 5 to 7 years. Infants with congenital malformations or missing data for GA, sex, or place of birth were excluded.

### Ethics approval

Research ethical and regulatory approval as part of the neoWONDER study (reference 21/EM/0130)^14^ including Confidentiality Advisory Group approval to use identifiers for linkage purposes without consent (CAG reference 21/CAG/0081, Section 251 of the NHS Act 2006). No NNU opt-out requests were received.

### Definitions of variables

#### Outcomes

All outcomes are binary measures of whether a child met the expected education level at three time points (at 5 years, 6 years, 7 years), ascertained from statutory assessments in England.

#### Age 5 Reception: Early Years Foundation Stage Profile (EYFSP)

At the end of the final year of reception (akin to pre-school), teachers assess development across seven domains: communication and language (CL), physical development (PHY), personal, social, and emotional development (PSE), literacy (LIT), mathematics (MAT), understanding the world (UTW), and expressive arts and design (EXP). A ‘good level of development’ (GLD) was defined as achieving at least the expected level in CL, PHY, PSE, LIT, and MAT.^15^ We used EYFSP as a measure of school readiness because these domains reflect the skills and behaviours that promote successful engagement in the early years of formal schooling.^16^

#### Age 6, Key Stage 1 (KS1) phonics screening check

During year 1 of formal schooling, teachers assess phonics using a 40-word test to assess reading ability. Pupils scoring ≥32/40 meet the expected level.^17^

#### Age 7, End of KS1 assessments

Binary variables are derived to denote whether children meet the expected level in national tests of reading, writing, mathematics, and science.

For KS1 assessments, the outcome was expressed as ‘not meeting the expected level’ at each time point because it indicates children who were likely to require additional support.^18,19^

#### Exposures

For descriptive analyses, we reported outcomes stratified by GA groups: 23-26 and 27-31 completed weeks.^20^ The Index of Multiple Deprivation (IMD) was derived from the maternal postcode recorded in the NNRD at birth.^21^ Maternal postcodes were linked to UK Office for National Statistics Lower Super Output Areas to assign IMD scores, which were grouped into national deciles (1 = most deprived, 10 = least deprived).

#### Covariates

PTB-RFs included maternal smoking, age, gestational diabetes, and hypertensive disorders, and the following neonatal characteristics: sex, birth month, birthweight, multiplicity, mode of delivery, co-morbidities of preterm birth (retinopathy of prematurity [ROP], necrotising enterocolitis [NEC], sepsis, bronchopulmonary dysplasia [BPD)], brain injuries (cystic periventricular leukomalacia [cPVL], all-grade intraventricular haemorrhage [IVH], hydrocephalus, and porencephalic cyst), exposure to ante-and post-natal corticosteroids, and nutrition at discharge (see Table S1 for definitions).

Socioeconomic and demographic variables from the NPD included child ethnicity and major language group. The Income Deprivation Affecting Children Index (IDACI) was recorded at age 5 years. The IDACI ranks neighbourhoods according to the proportion of children under 16 living in low-income households, expressed as deciles.^21^ Eligibility for free school meals (FSM) at age 5 years was included as a family-level proxy for household income.

## Statistical analysis

### Descriptive analyses

We report participant characteristics for the study cohort with and without a linked education record (Table 1). Frequencies (%) and median (IQR) are presented for categorical and continuous variables, respectively. For each outcome, we plotted the prevalence (95% CI) of not meeting the expected level of attainment across IMD, IDACI, and FSM stratified by 23-26 weeks’ and 27-31 weeks’ gestation. We used the Cochran–Armitage test to evaluate trends. We report outcomes for the study cohort alongside DfE national data for the same epoch.^15,17^ We plotted the ten most common longitudinal individual-attainment trajectories.

**Table 1:** Characteristics of children with and without a linked NNRD-NPD record. Frequencies (%) and median (IQR) are presented for categorical and continuous variables, respectively. *Frequencies for cystic periventricular leukomalacia and hydrocephalus have been suppressed to minimise risk of secondary disclosure when cell counts describe <10 individuals.

|  | Not linked | Linked | Total |
| --- | --- | --- | --- |
| **N** | **1,029 (6.1%)** | **15,857 (93.9%)** | **16,886 (100.0%)** |
| **Gestational age (completed weeks)** |  |  |  |
| 23-24 | 44 (4.3%) | 651 (4.1%) | 677 (4.1%) |
| 25 | 49 (4.8%) | 748 (4.7%) | 797 (4.7%) |
| 26 | 80 (7.8%) | 1,196 (7.5%) | 1,276 (7.6%) |
| 27 | 105 (10.2%) | 1,546 (9.7%) | 1,651 (9.8%) |
| 28 | 125 (12.1%) | 2,091 (13.2%) | 2,216 (13.1%) |
| 29 | 160 (15.5%) | 2,399 (15.1%) | 2,559 (15.2%) |
| 30 | 215 (20.9%) | 3,068 (19.3%) | 3,283 (19.4%) |
| 31 | 251 (24.4%) | 4,158 (26.2%) | 4,409 (26.1%) |
| **Child characteristics** |  |  |  |
| Birth-weight z-score | -0.211 (-0.82 to 0.30) | -0.21 (-0.81 to 0.31) | -0.21 (-0.81 to 0.31) |
| Small for gestational age | 142 (13.8%) | 2,135 (13.5%) | 2,277 (13.5%) |
| **Birth multiplicity** |  |  |  |
| Singleton | 715 (69.5%) | 11,582 (73.1%) | 12,297 (72.8%) |
| Multiple | 314 (30.5%) | 4,270 (26.9%) | 4,584 (27.2%) |
| **Sex at birth** |  |  |  |
| Female | 460 (44.7%) | 7,408 (46.7%) | 7,868 (46.6%) |
| Male | 569 (55.3%) | 8,449 (53.3%) | 9,018 (53.4%) |
| **Socio-economic status** |  |  |  |
| IMD decile at birth: 1 Most deprived | 104 (10.9%) | 2,847 (18.5%) | 2,951 (18.0%) |
| 2 | 118 (12.3%) | 2,285 (14.8%) | 2,403 (14.7%) |
| 3 | 105 (11.0%) | 1,939 (12.6%) | 2,044 (12.5%) |
| 4 | 103 (10.8%) | 1,597 (10.4%) | 1,700 (10.4%) |
| 5 | 70 (7.3%) | 1,520 (9.9%) | 1,590 (9.7%) |
| 6 | 98 (10.2%) | 1,241 (8.0%) | 1,339 (8.2%) |
| 7 | 85 (8.9%) | 1,144 (7.4%) | 1,229 (7.5%) |
| 8 | 95 (9.9%) | 1,045 (6.8%) | 1,140 (7.0%) |
| 9 | 96 (10.0%) | 916 (5.9%) | 1,012 (6.2%) |
| 10 Least deprived | 83 (8.7%) | 885 (5.7%) | 968 (5.9%) |
| **Maternal characteristics** |  |  |  |
| Mother's age at birth (years) | 33 (28 to 36) | 30 (25 to 34) | 30 (25 to 35) |
| Maternal ethnicity |  |  |  |
| White | 661 (67.9%) | 11,560 (76.9%) | 12,221 (76.3%) |
| Asian | 148 (15.2%) | 1,612 (10.7%) | 1,760 (11.0%) |
| Black | 98 (10.1%) | 1,362 (9.1%) | 1,460 (9.1%) |
| Mixed | 13 (1.3%) | 246 (1.6%) | 259 (1.6%) |
| Other | 53 (5.4%) | 254 (1.7%) | 307 (1.9%) |
| Maternal smoking at birth | 79 (7.7%) | 2,906 (18.3%) | 2,985 (17.7%) |
| Infant of a diabetic mother | 24 (2.3%) | 457 (2.9%) | 481 (2.8%) |
| Hypertensive disorders of pregnancy | 140 (13.6%) | 2,037 (12.8%) | 2,177 (12.9%) |
| **Interventions** |  |  |  |
| Complete course of ante-natal corticosteroids | 701 (73.2%) | 10,458 (70.0%) | 11,159 (70.2%) |
| Post-natal corticosteroids | 58 (5.8%) | 1,095 (7.1%) | 1,153 (7.0%) |
| **Neonatal morbidities** |  |  |  |
| Sepsis | 49 (4.8%) | 785 (5.0%) | 834 (4.9%) |
| Necrotising enterocolitis | 17 (1.7%) | 362 (2.3%) | 379 (2.2%) |
| Retinopathy of prematurity | 18 (1.7%) | 462 (2.9%) | 480 (2.8%) |
| Bronchopulmonary dysplasia | 264 (26.0%) | 4,195 (26.9%) | 4,459 (26.9%) |
| **Neonatal brain injuries** |  |  |  |
| Cystic periventricular leukomalacia* | <10 (0.9%) | SUPP (1.6%) | SUPP (1.6%) |
| Intraventricular haemorrhage (IVH) |  |  |  |
| No IVH | 902 (87.7%) | 13,775 (86.9%) | 14,677 (86.9%) |
| IVH grade I-II | 91 (8.8%) | 1,648 (10.4%) | 1,739 (10.3%) |
| IVH grade III-IV | 36 (3.5%) | 434 (2.7%) | 470 (2.8%) |
| Porencephalic cyst | 15 (1.5%) | 202 (1.3%) | 217 (1.3%) |
| Hydrocephalus | <10 (0.5%) | SUPP (1.1%) | SUPP (1.0%) |
| **Nutrition at discharge** |  |  |  |
| Enteral milk at discharge |  |  |  |
| Breast | 458 (47.6%) | 4,257 (28.8%) | 4,715 (29.9%) |
| Formula | 204 (21.2%) | 5,946 (40.2%) | 6,150 (39.0%) |
| Other | 57 (5.9%) | 1,497 (10.1%) | 1,554 (9.9%) |
| Mixed | 243 (25.3%) | 3,091 (20.9%) | 3,334 (21.2%) |

### Regression analysis

Overall, 0 8% of all covariate values were missing in the data. Missingness was addressed through multiple imputation using chained equations (see Supplement methods and Table S2). We fitted cross-sectional logistic regression models using generalised estimating equations (GEE) to develop functional statistical relationships between covariates and each outcome. An exchangeable correlation structure was specified to account for within-mother correlation among multiple births, assuming equal correlation between siblings.^22^ Model estimation employed a binomial distribution with a logit link and robust standard errors. We assessed multicollinearity using the variance inflation factor (VIF), ensuring that values did not exceed 5. Children with complete data on all outcomes and maternal ID to identify within-family clustering were included in the GEE models.

IMD quintiles were used in three models to highlight broader gradients of deprivation in line with previous studies: 1) crude associations of each covariate with each outcome; 2) a model to test the interaction between GA and IMD, which was assessed by testing statistical significance (at p=0.05 level) of the interaction term and by plotting predicted probabilities of outcomes at mean values of covariates values stratified by GA across levels of IMD; and 3) fully adjusted models including all covariates described above. To avoid collinearity between ethnicity, language, and socioeconomic variables (Figure S1), our primary regression analyses retained IMD, FSM eligibility, and language, while omitting IDACI and ethnicity.

In a sensitivity analysis, we substituted ethnicity for language to evaluate its impact on outcomes. We report results as odds ratios (OR) with 95% confidence intervals (CI). A CI that did not cross 1 was considered statistically significant at 5%. We evaluated out of sample predictive performance of the fully adjusted models (see Supplemental methods, ‘Predictive performance of the fully adjusted models‘). Analyses were conducted using Stata, version 18 0 and R.

## Results

### Study population

Of 23,862 infants in the NNRD, 17,245 were identified by NHS Digital for linkage and 6,617 were not identified due to missing or incomplete NHS numbers. There were no major differences between groups with and without sufficient linkage identifiers (Table S3). Of those identified, 15,857 (92%) had an education record in the NPD (Figure 1). Among these, 16 3% were born 23 to 26 weeks and 83 7% born 27 to 31 weeks; 53 3% were male, 26.9% were multiple-born, and 76 9% were born to White mothers (Table 1). There were no differences in the distribution of GA, sex, or neonatal morbidities between linked and non-linked groups; however, the linked group were more likely to be of White ethnicity (76 9% vs 67 9%) and from the most deprived IMD decile (18 5% versus 10 9%).

**Figure 1:**
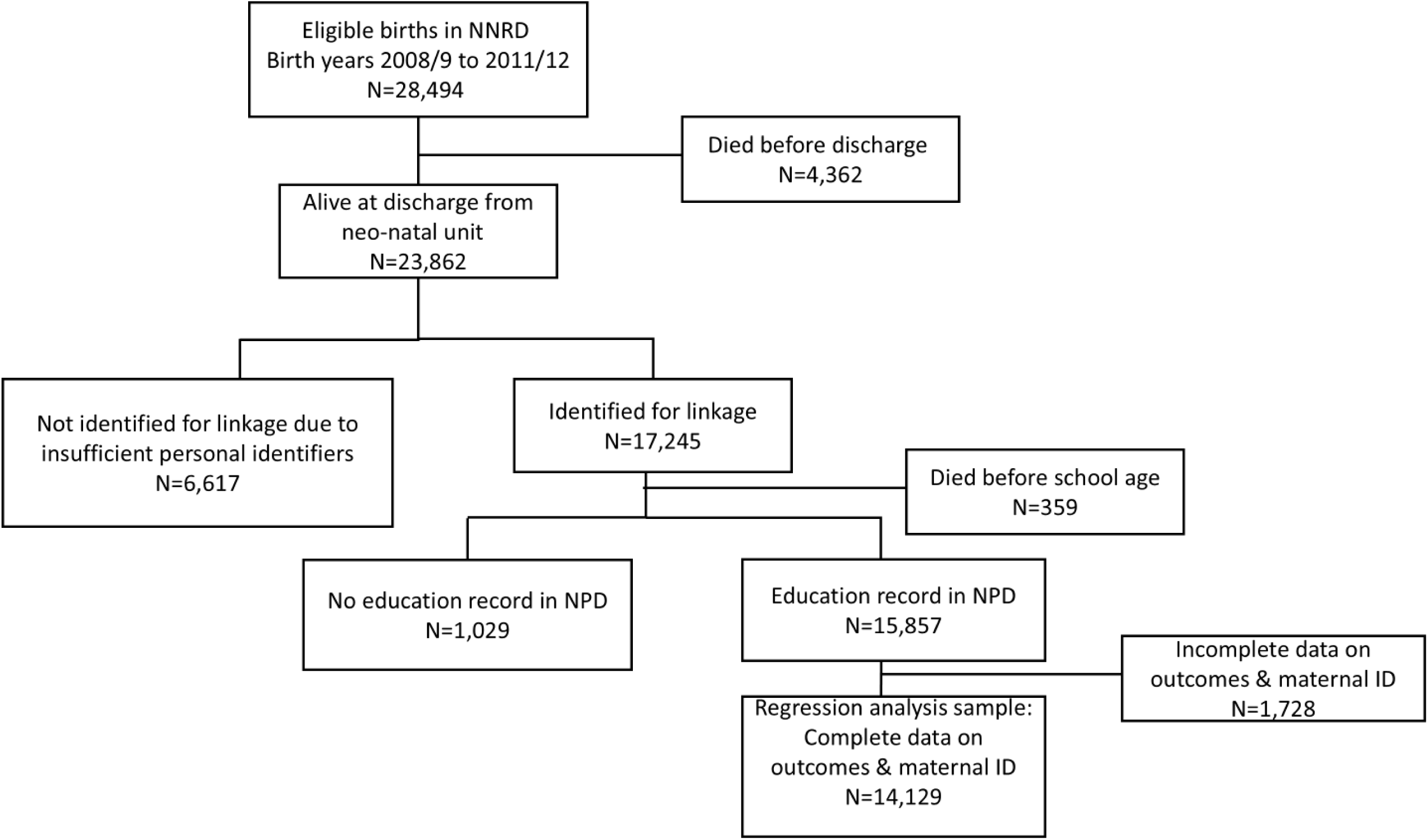
Participant flow of cohort included in the study born between. 2008**/09 to** 2011**/12 in England**

### Trends in educational attainment of children born at <32 weeks at 5, 6, and 7 years old, and comparison to the whole population

Table S4 shows the annual proportion of the cohort born in 2008/2009 to 2011/2012 who did not meet the expected level in each outcome, and Figure 2 summarises the data alongside national population results for the same epoch. For preterm born children at 5-6 years, a higher proportion did not meet the expected level compared to the national average (EYFSP 56 6% vs 35 5%; phonics 20 7% vs 8 8%); at 7 years, around half did not meet expected levels in writing (51 8%), maths (48 1%), and approximately 40% in reading and science. Compared with the national average,^15,17^ the percentage point differences were 21% (EYFSP), 12% (phonics), 17% (KS1 reading), 20% (KS1 writing), 23% (KS1 maths), and 18% (KS1 science), Figure 2. Children born <32 weeks constituted 1-2% of the national comparator data.^23^

**Figure 2:**
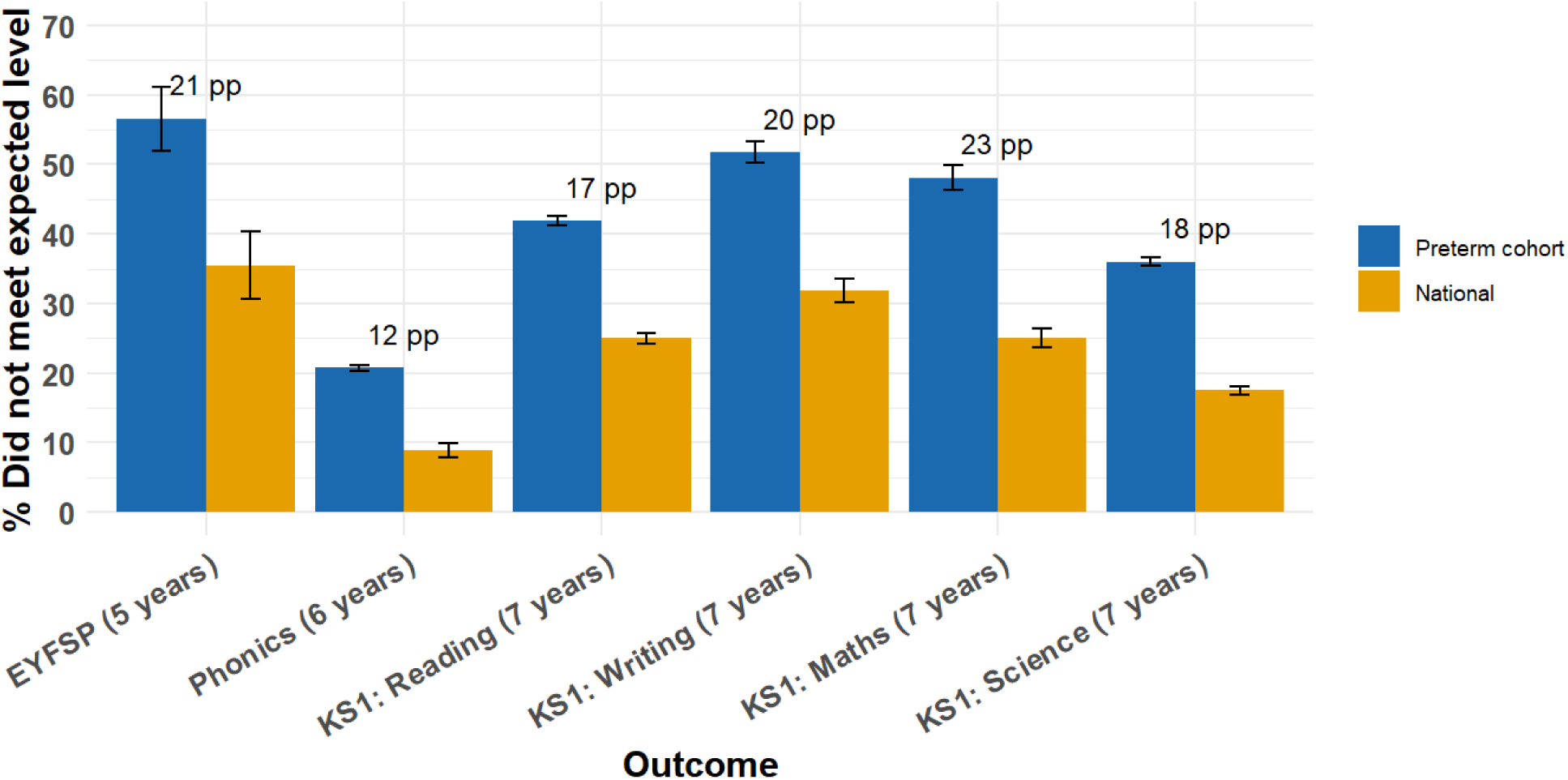
Distribution of children who did not meet expected levels for each outcome in the cohort and the whole national population. Comparison between educational outcome measures in the cohort and published national statistics for all children who were at school and undertook the assessments during the same time-period. We derived the % who did not meet the expected level from annual figures published in reports by the Department for Education.^15,17^ The bars represent the mean and SD over four birth years (2008/09 to 2011/12). Figures show the percentage point difference between preterm cohort and national attainment.

### Associations between gestational age and educational attainment

There was a dose-response relationship between GA and educational attainment: each decreasing week of gestation from 31 weeks was associated with an increased proportion of children not achieving expected levels across all outcomes (Figure 3, Table S5). For EYFSP and KS1 reading, maths, science, and writing domains, approximately 70–90% born at 23 weeks did not reach expected standards, compared with around 30–50% at 31 weeks.

**Figure 3:**
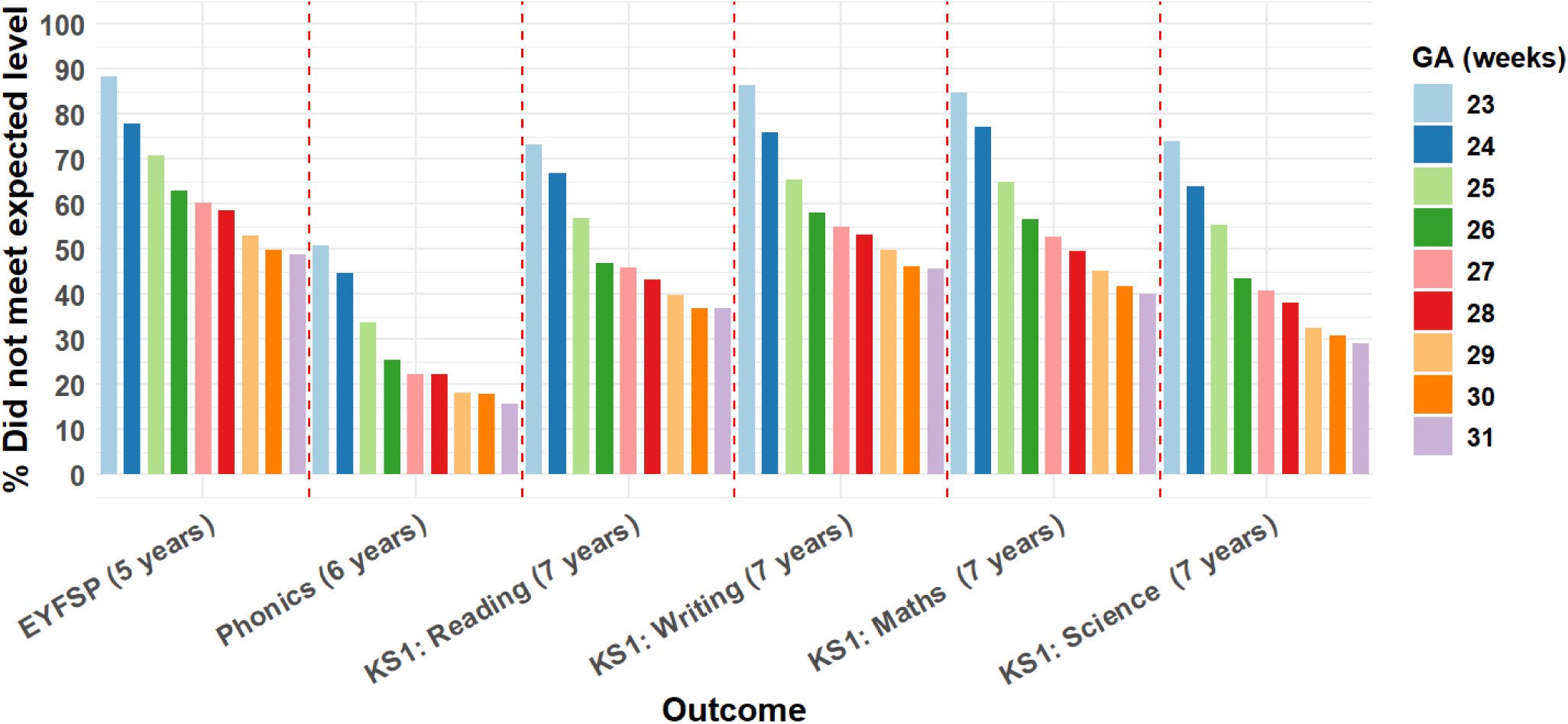
Distribution of children who did not meet the expected level across outcomes stratified by gestational age (GA) Proportion in the overall cohort that did not meet expected level: EYFSP 8,602/15,528 (55.9%); Phonics 3,226/15,504 (20.8%); KS1 Reading 6,354/15,141 (42%); KS1 Writing 7,789/15,141 (51.4%); KS1 Maths 7,216/15,140 (47.7%); KS1 Science 5,449/15,141 (36%)

### Associations between socioeconomic status and educational attainment

The proportion of children who did not meet expected levels declined progressively with higher SES at birth (Figure 4). The gradients across IMD levels were parallel, indicating similar effects of deprivation across GA groups. Among children born in the least deprived IMD decile, 52% born at 23-27 weeks and 42% born at 28-31 weeks did not achieve the expected level at EYFSP. For the most deprived decile, the corresponding percentages were higher at 75% and 60%, respectively. These trends were consistent for FSM and IDACI at 5 years (Figures S2 and S3).

**Figure 4:**
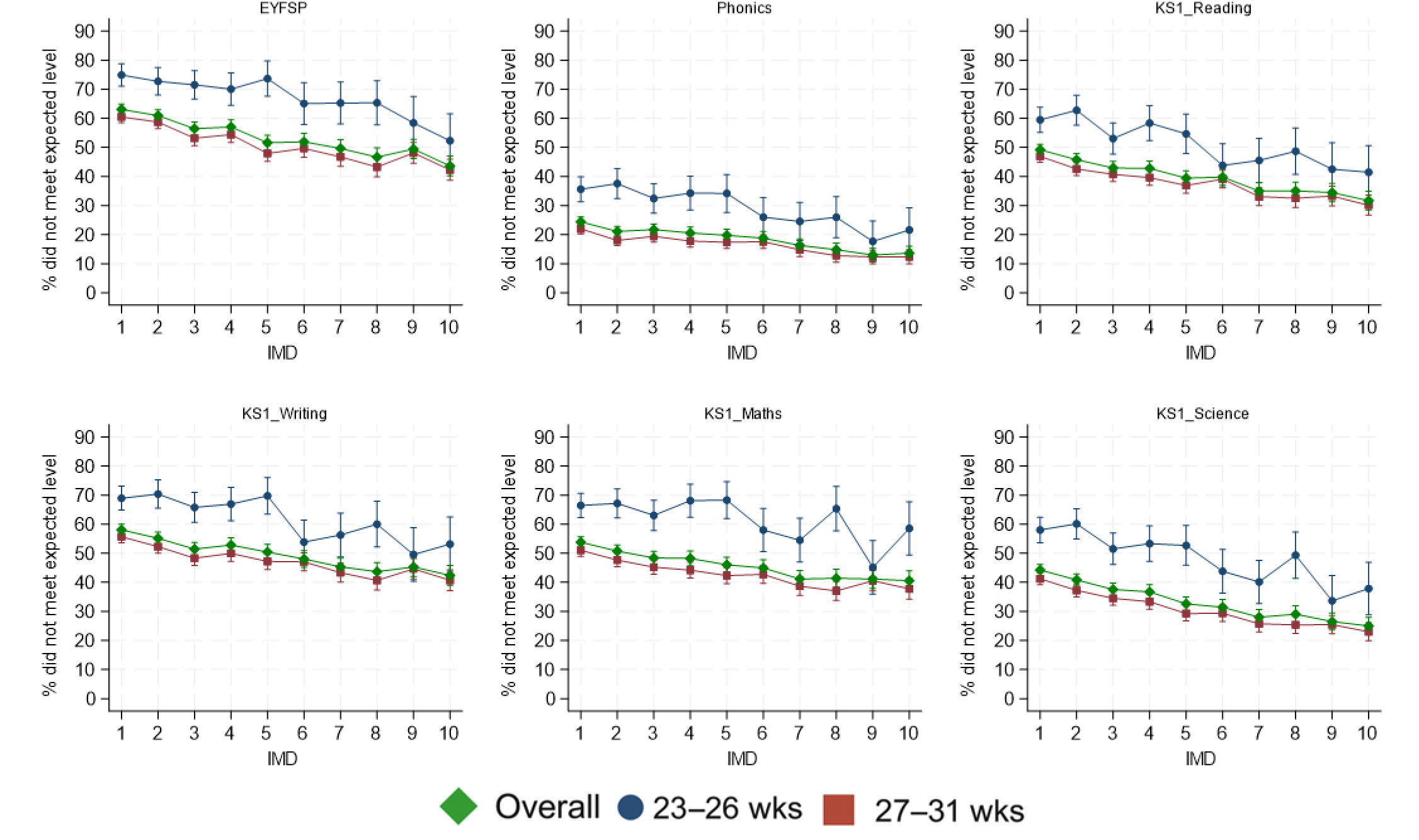
Proportion of children who did not meet the expected level in outcomes by socio-economic status at birth (Index of Multiple Deprivation) stratified by gestational age category (23-26 weeks and 27-31 weeks) and the overall cohort. Trend tested using Cochran–Armitage test. P<0.001 for every line. IMD 1= most deprived.

### Longitudinal individual-attainment trajectories

Figure 5 shows the relative frequencies of the top ten most frequently occurring within-individual attainment sequence across all outcomes stratified by degree of prematurity. Among children born at 23-26 weeks, persistent under-performance in all outcomes was the most frequent attainment pattern (29 5%), and children were 1 7 times more likely to not meet expected levels in at least five outcomes compared with children born at 27-32 weeks (43 7% versus 26%). The most frequently occurring sequence in the 27-31 week group was meeting the expected level in all outcomes (35 9%), almost twice more than the 23-26 week group (21 6%).

**Figure 5:**
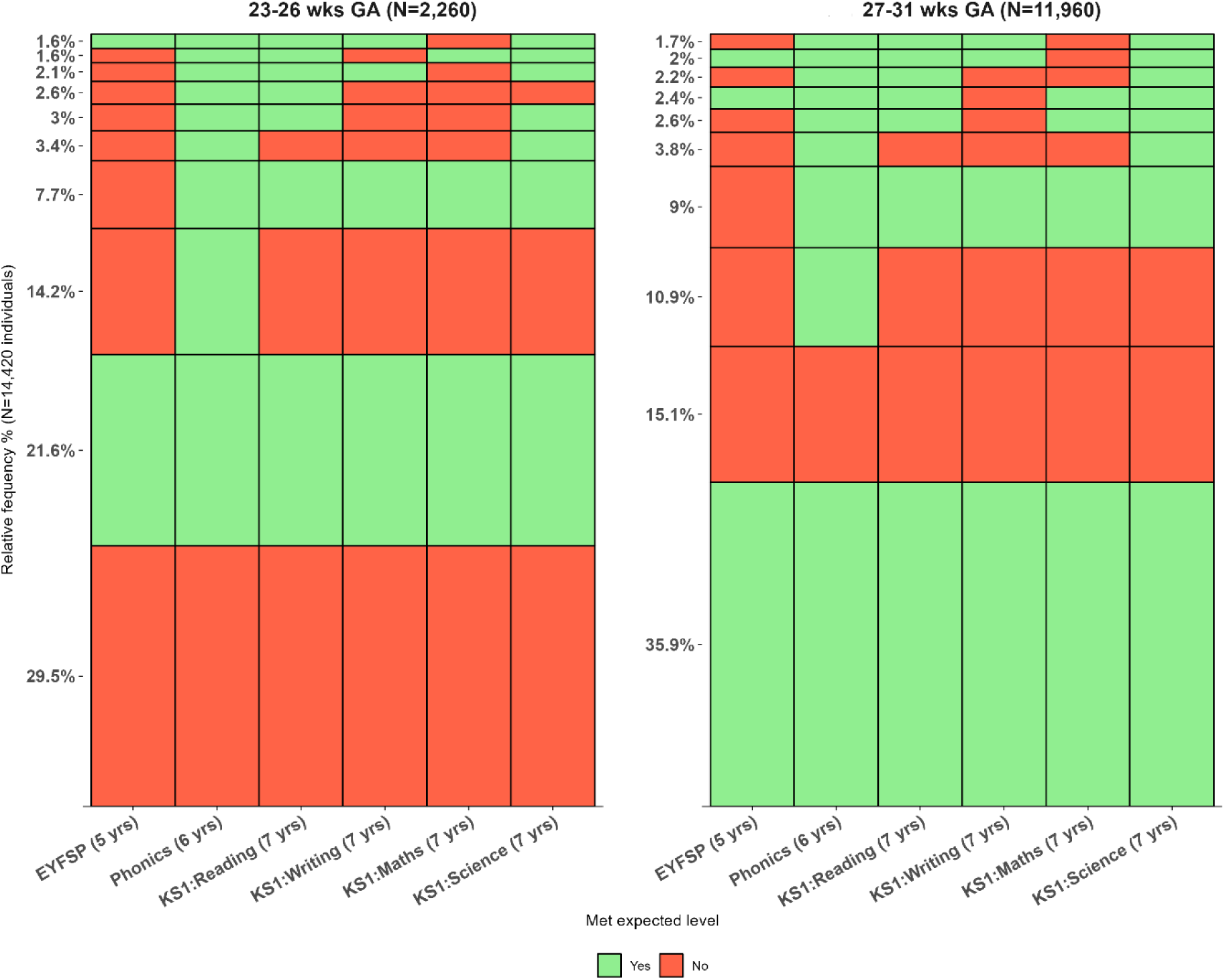
Sequence frequency plot to show the ten most frequently occurring longitudinal outcome patterns between age 5 and 7 years, stratified by gestational age category (23-26 weeks and 27-31 weeks) The y-axis represents the cumulative percentage of longitudinal sequences, and the bar widths are proportional to their frequencies. Rows are ordered by frequency of the pattern.

### Factors associated with educational attainment over time in mutually adjusted models

#### Impact of gestational age and other infant characteristics

Table 2 and Figure 6 present the ORs and 95% CIs from the unadjusted and fully adjusted GEE regression models for EYFSP, phonics, and KS1 reading and maths. There were strong correlations across domains for KS1 7-year outcomes (*r*>0 8); the results for KS1 writing and science are presented in Supplementary Table S6 and Figure S4.

**Figure 6:**
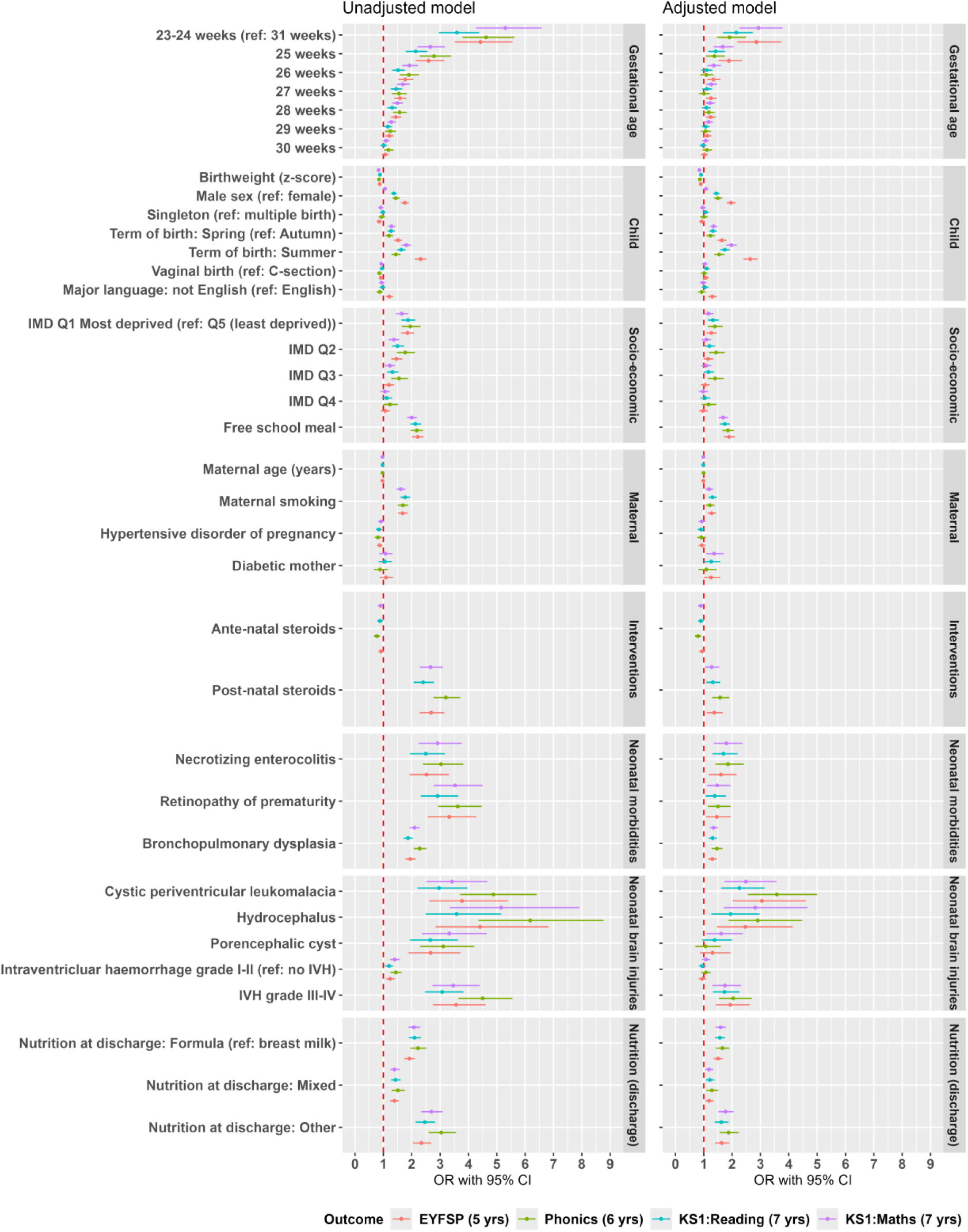
Forest plot to show the unadjusted and fully adjusted associations between socio-economic and preterm birth factors with educational outcomes (not meeting the expected level in EYFSP, Phonics, KS1 reading, or maths) Odds ratios and 95% confidence intervals derived from GEE regression models.

**Table 2:**
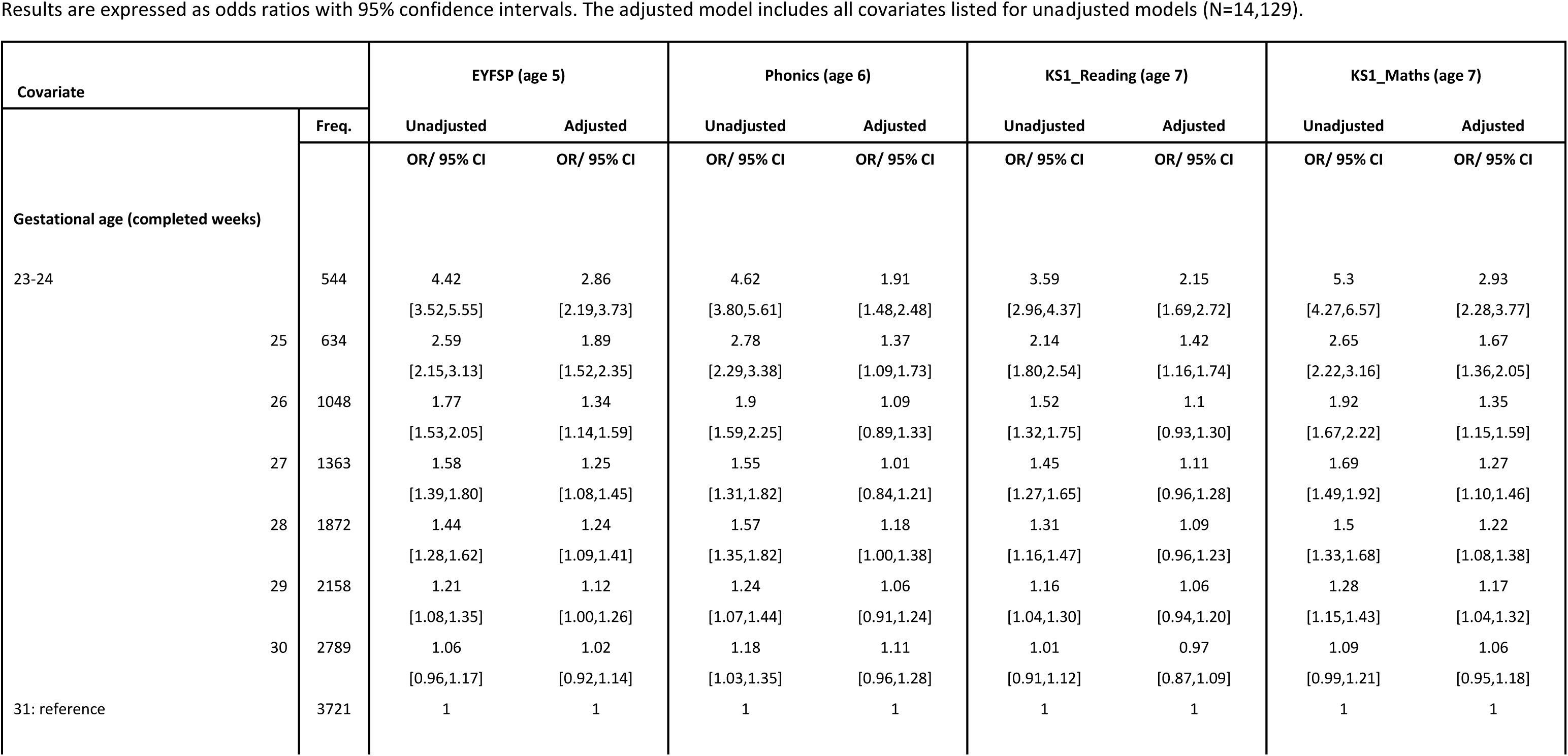
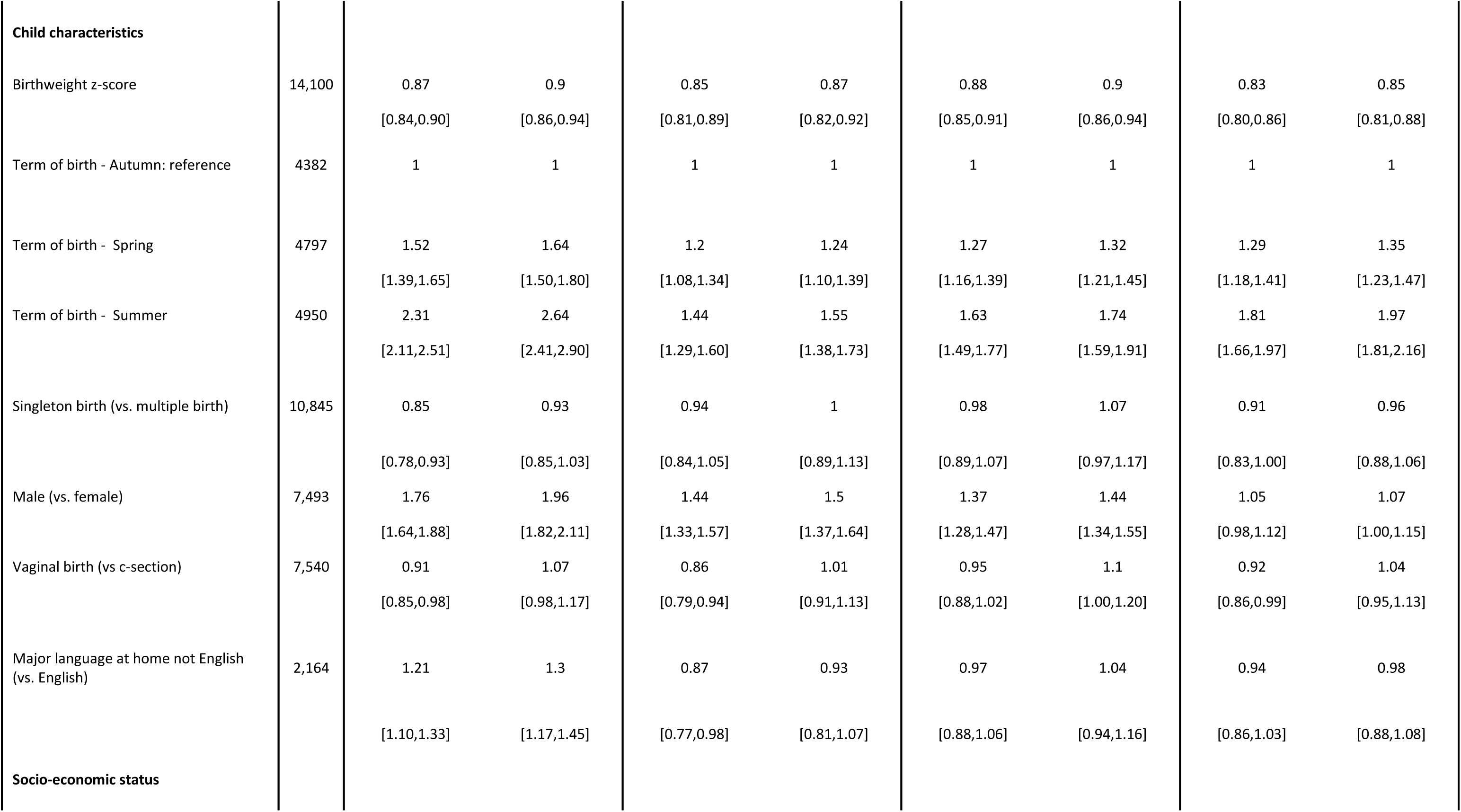
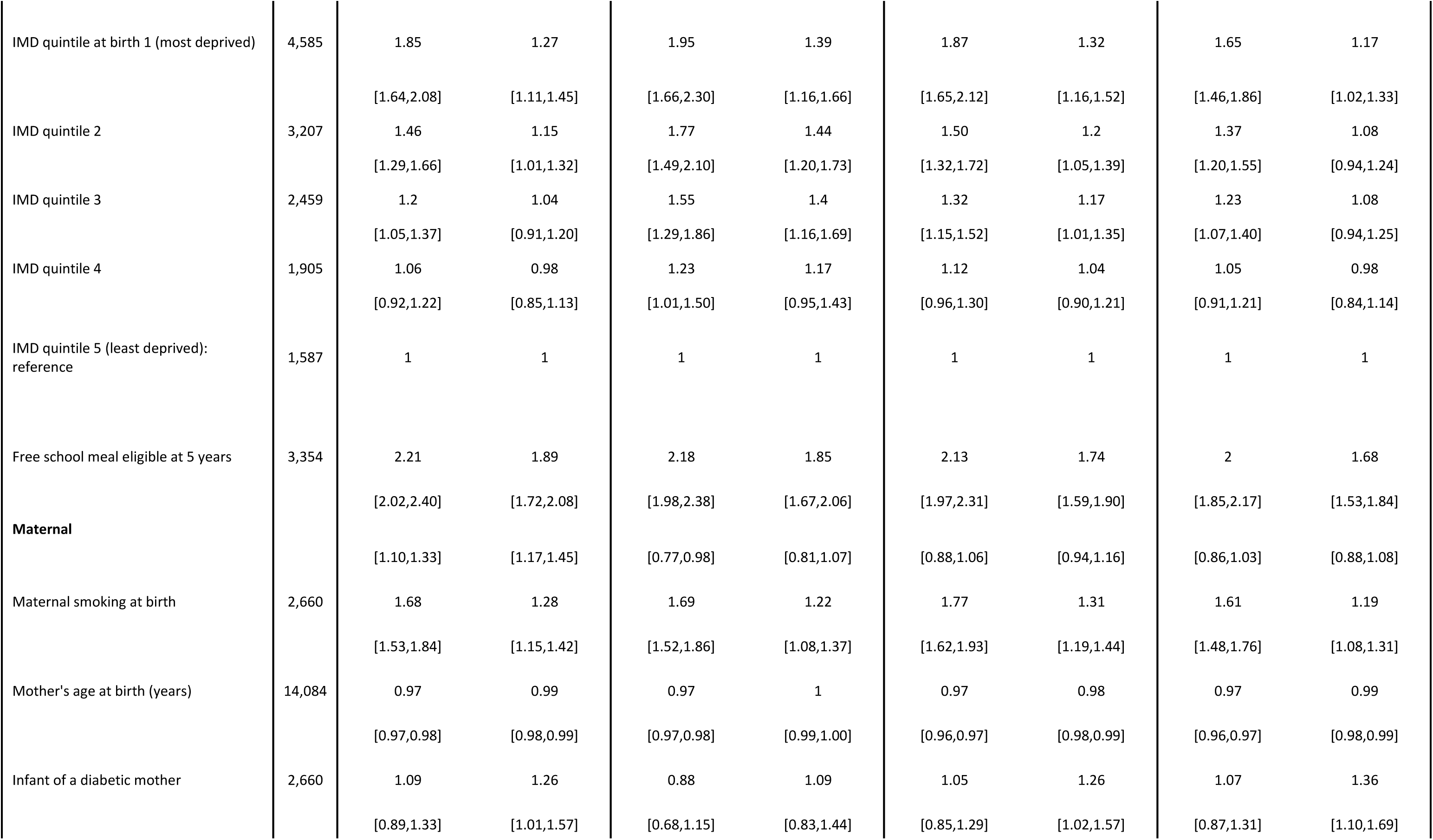
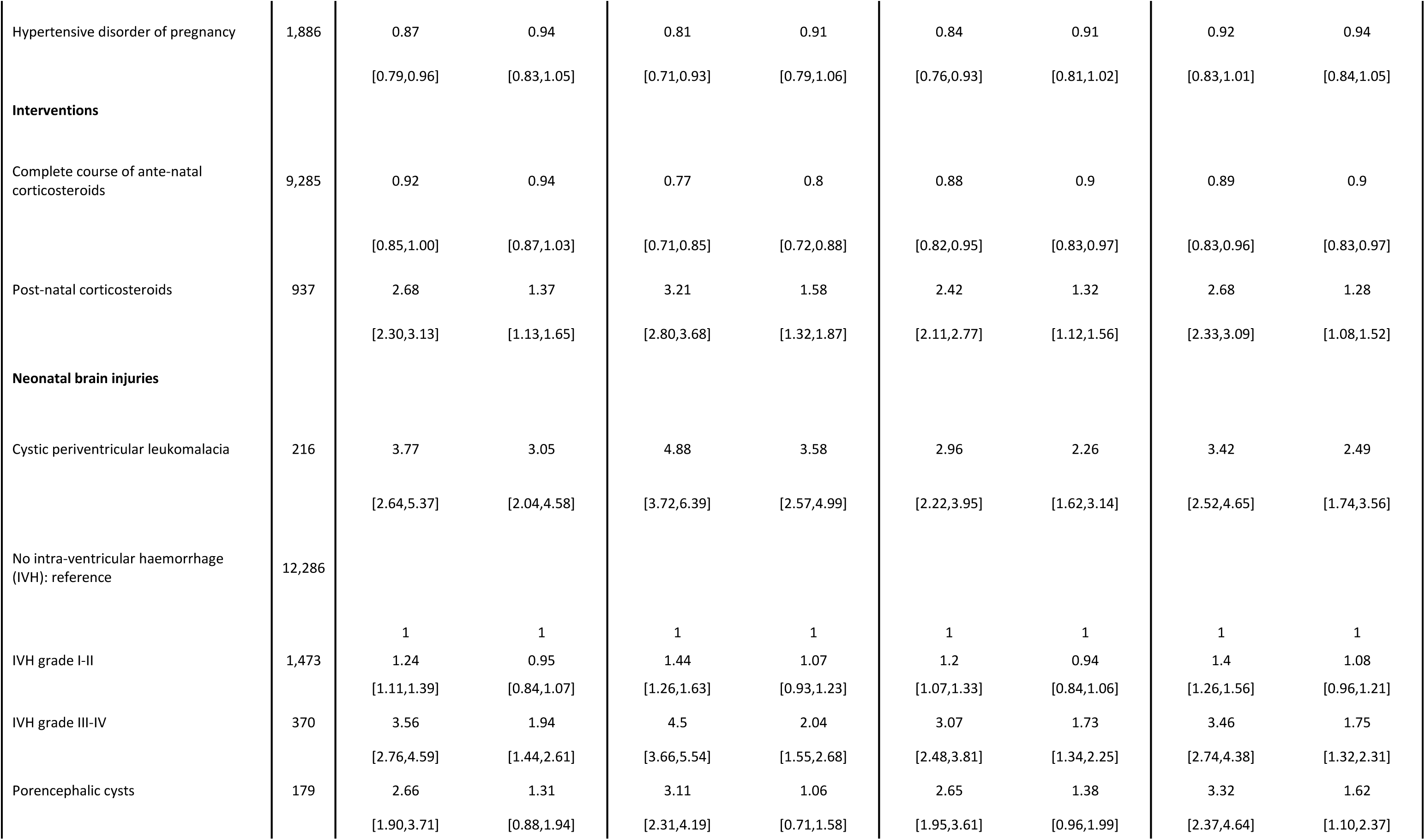
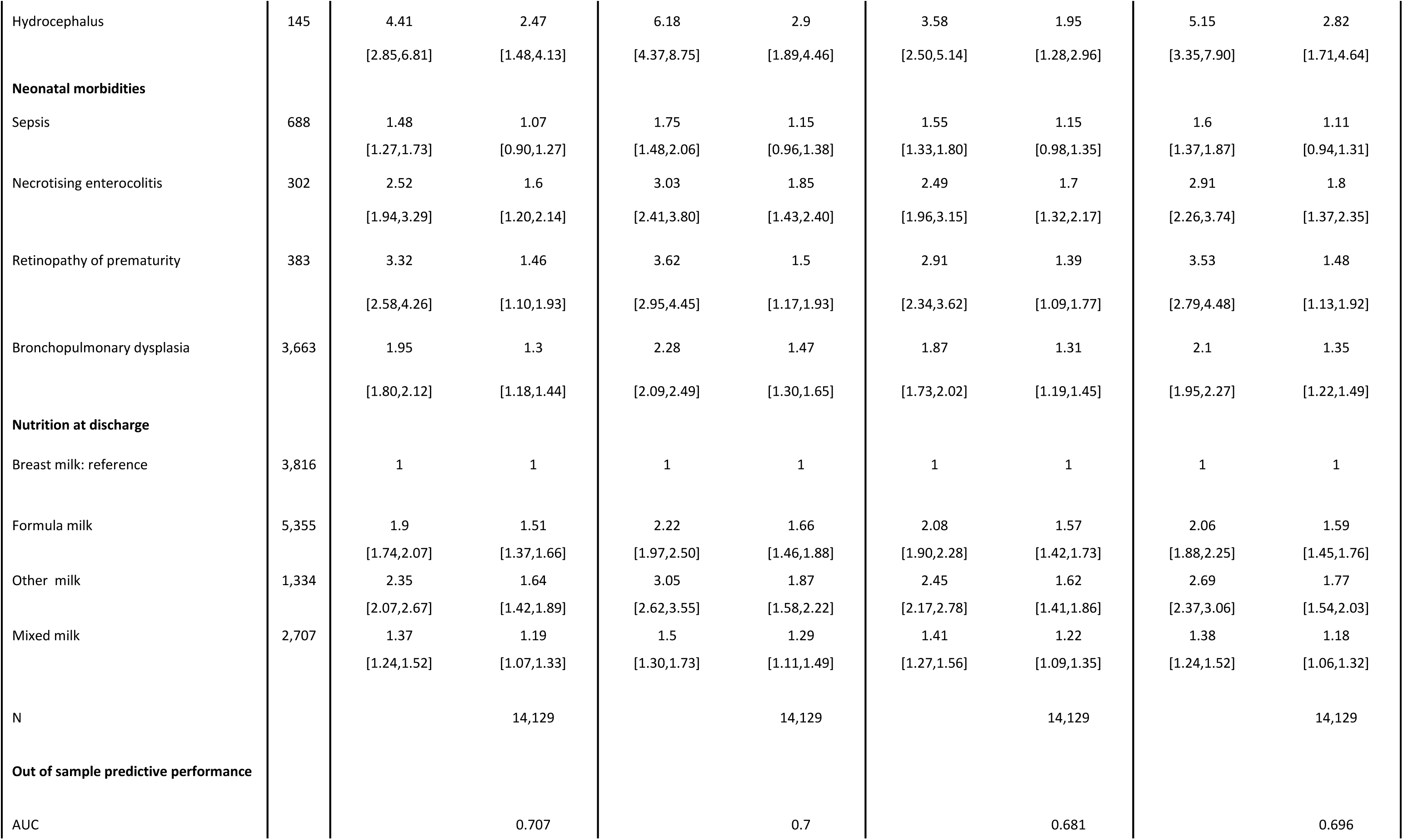
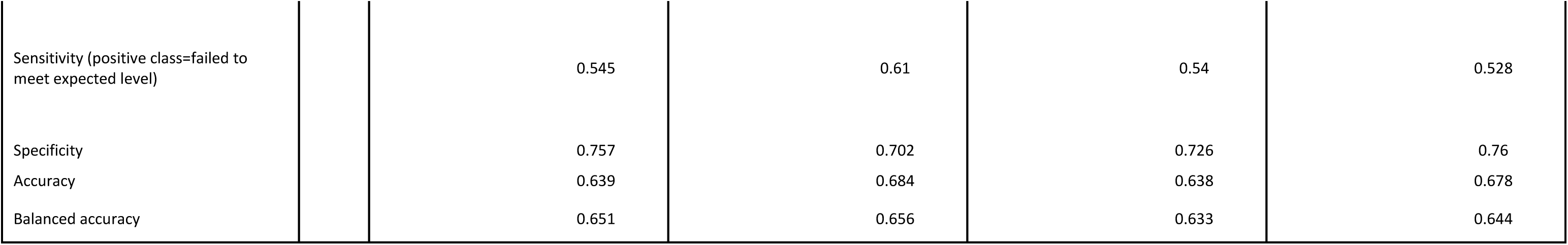
Unadjusted and adjusted GEE regression models for the association between socio-economic and preterm-birth factors with educational outcomes for children born 2008/09 to 2011/12 in England. Results are expressed as odds ratios with 95% confidence intervals. The adjusted model includes all covariates listed for unadjusted models (N=14,129).

14,129 children with complete data on all outcomes and maternal ID to identify clustered sibling data were included in the models. The dose-response relationship between GA and attainment persisted after adjustment, particularly for the most preterm infants, although effect sizes were attenuated. After adjustment, children born at 23–24 weeks were two to three times more likely to not achieve the expected level compared to those born at 31 weeks (EYFSP: adjusted odds ratio (aOR) 2.86, 95% CI 2.19-3.73) (Table 2). Higher birthweight and female sex were protective for attainment although the gap between male and female children narrowed by age 7 years, especially for maths (EYFSP: (aOR) 1 96, 95% CI 1 82–2 11; maths: aOR 1 07, 95% CI 1 00–1 15) (Table 2). Summer-born children, who enter school a year earlier than Autumn-born children, were nearly three times more likely to not meet expectations at EYFSP (aOR 2 64, 95% CI 2 41–2 90). Attainment did not differ between singletons and multiples.

#### Impact of maternal characteristics

Smoking during pregnancy increased the odds of under-attainment by 19–37% across outcomes (Table 2). Increasing maternal age was protective, with each additional year associated with a 1–2% reduction in under-attainment. The effect of maternal diabetes increased after adjustment; however, hypertensive disorders had no statistical association with education attainment.

#### Impact of care and interventions in the perinatal period

A complete antenatal corticosteroid course administered to the mother was protective across all attainment timepoints and domains, whereas postnatal corticosteroids were consistently associated with elevated odds of not meeting the expected level of attainment (EYFSP: aOR, 1 37, 95% CI 1 13-1 65; KS1 maths: aOR 1 28, 95% CI 1 08-1 52). Lack of exclusive breastfeeding predicted poorer outcomes: mixed feeding had a lower association than exclusive formula feeding (EYFSP: aOR 1 19, 95% CI 1 07–1 33 versus aOR 1 51, 95% CI 1 37–1 66, respectively).

#### Preterm complications/co-morbidities

Of all covariates, neonatal brain injuries had the strongest associations with educational outcomes, particularly PVL (EYFSP: aOR 3 05, 95% CI 2 04–4 58) and hydrocephalus (EYFSP: aOR 2 47, 95% CI 1 48–4 13). PVL demonstrated the least attenuation after adjustment compared with other preterm complications/co-morbidities and had an effect size on par with being born at 23-24 weeks GA. Porencephalic cysts was associated with outcomes at KS1 but not earlier assessments. Infants with NEC, treated ROP, and BPD were linked to 1 5–2-fold higher odds of poor outcomes, while no association was found with sepsis.

### Socioeconomic and environmental exposures

Socioeconomic disadvantage had a negative association with attainment at all ages. In adjusted models, children born in the most deprived quintile of neighbourhoods were 20–40% more likely to underachieve than those in the least deprived quintile (EYFSP: aOR 1.27, 95% CI 1.11–1.45), and children eligible for FSM were almost twice as likely to have poor outcomes, a magnitude on par with severe intraventricular haemorrhage (grade III–IV), Table 2. Children whose primary language was not English were more likely to underachieve at EYFSP, though this difference was not present at KS1.

There was no statistical interaction between deprivation and GA; the probability of poor attainment increased steadily with higher deprivation in all GA groups, confirming that deprivation exerts a comparable negative influence regardless of degree of prematurity (Figure S5).

Overall, findings were consistent for KS1 science and writing (Figure S4, Table S6). Models showed low multicollinearity (VIF 1 28), moderate discrimination (AUC≈0.7) and classification performance (balanced accuracy of 0 63-0 66). Sensitivity analyses substituting major language spoken at home with ethnicity showed no consistent effect of ethnicity (Table S7). Effects of IMD and FSM were stable to its inclusion.

## Discussion

In this large, population-based cohort study of preterm infants born in England before 32 weeks’ gestation between 2008 and 2012, there was a substantially elevated risk of not being ready for school at 5 years and of not meeting expected attainment levels in statutory tests at 6 and 7 years. There was a dose effect of prematurity: only a fifth of children born at 23-26 weeks and a third born at 27-31 weeks met expected levels of attainment at all time points. The data highlight the additive impact of socioeconomic deprivation in shaping the educational outcomes of children born very preterm. Those born in the most deprived quintile of neighbourhoods were 20–40% more likely not to meet expected levels of attainment than those born in the least deprived quintile, even after accounting for low GA and medical complications. Strikingly, FSM eligibility at age 5 years, a proxy for family income, was associated with an almost doubling risk of under-attainment, which is an effect size comparable to that of severe neonatal brain injury. Our findings corroborate education research showing that deprivation impacts pre-school neurodevelopmental outcome,^24,25^ and cognitive neuroscience research showing that inequalities associate with differences in brain structure and connectivity, particularly in regions that support language, executive function, and memory, among preterm children.^10,26^

The data indicate that social deprivation and PTB constitute a ‘double burden’. These adverse effects may be modifiable through family-level interventions and social and public policy designed to alleviate deprivation. This result is particularly important given that one-third of UK children live in relative poverty.^27^ In addition, there is a need for continued clinical and research efforts to reduce severe neonatal brain injuries, NEC, BPD, and ROP, which all predicted low attainment. Whilst postnatal corticosteroid exposure was associated with low attainment, further work is required to explore whether the exposure is a proxy of illness severity or due to a direct effect on the developing brain.

Although some of the PTB-RFs that are associated with low attainment are immutable (GA, male sex), others are potentially modifiable with targeted interventions, such as increasing breastfeeding rates during NNU care, improving antenatal corticosteroid use, and reducing maternal smoking. Studies are required to determine whether children born in the summer months could benefit from delayed school entry or targeted support, and whether very preterm boys and children growing up in deprived circumstances may have additional support needs. Ensuring that children born preterm with additional risk factors are supported for school readiness could confer life-course and societal benefits, including higher educational achievement and improved employability and income.^28,29^

### Strengths and limitations

The study has several strengths. The use of a contemporary, socioeconomically and ethnically diverse, population-level cohort ensures high representativeness and generalisability to infants with access to modern neonatal intensive care. Furthermore, linkage with the NNRD ensured that we had a rich set of perinatal clinical variables. Socioeconomic status was triangulated from multiple data sources (IMD at birth, IDACI, and free school meal eligibility) to capture both neighbourhood-and family-level proxies of early-life deprivation, enhancing the robustness of our findings. As the UK government routinely collects these indicators, the findings are likely to be tractable to policymakers. We used multiple domains of educational outcomes, enabling us to identify shared and outcome-specific associations. Finally, the cohort is considerably larger than those reported in previous linkage studies of birth and education data, so was powered to detect novel associations in very preterm infants.^18,19^

There are some limitations. First, 27 7% of eligible children were not linked due to missing or incomplete NHS number information. However, we did not identify biases, particularly socioeconomic, between groups with and without linked educational records. 1,029 (6%) children whom NHS Digital identified for linkage did not have an education record. Speculatively, this could include children who are home-schooled, in private or hospital schools, or have migrated since birth. Second, our adjusted models achieved a moderately discriminative AUC of 0 7. Omitted variable bias in the models may have resulted from unavailable data on important factors previously identified to increase school readiness, such as maternal education.^16^ Third, we could not adjust for pre-existing neurodevelopmental impairment, so unmeasured variation in pre-school functioning may confound the observed associations at school age.^24^ Finally, we did not have access to data from term-born controls, so the study may have been underpowered to detect an interaction effect of GA and deprivation. Due to differences in neonatal practice and in the assessment of educational outcomes, this study may have limitations in generalisability to LMIC settings. However, the findings suggest that maternal health behaviours, optimal use of antenatal corticosteroids, breast milk, and reducing co-morbidities are likely to be universally advantageous for improving developmental trajectories.^30^

## Future work

We have demonstrated the methodological feasibility of linking population neonatal health to education data. Previous work has shown that parents support data linkage between health and education data;^14^ continued follow-up of this cohort will enable evaluation of interventions, such as policy measures that reduce child deprivation. Identification of early life risk factors for low educational attainment will enhance research into the biological processes that embed preterm birth and social inequalities in child development.

## Conclusion

This longitudinal birth cohort study found that children born at <32 weeks’ gestation often do not meet expected attainment levels in statutory educational assessments at 5-7 years. Low GA was strongly predictive of underachievement, but socioeconomic deprivation and several modifiable early-life exposures and co-morbidities were identified. Policies that reduce maternal and child deprivation, alongside clinical and research strategies that improve maternal health and minimise the co-morbidities of preterm birth, could improve school performance and potentially attenuate the life course impacts of being born too soon.

## Data Availability

The data underlying this study cannot be shared publicly due to data-sharing agreements with NHS Digital and the Department for Education.

## Acknowledgements

The work was funded by UKRI Medical Research Council programme grant (MR/X003434/1) awarded to JPB. We wish to acknowledge all neonatal units who contribute data to the NNRD, known collectively as the UK Neonatal collaborative (www.imperial.ac.uk/neonatal-data-analysis-unit/neonatal-data-analysis-unit/list-of-national-neonatal-units/).

Infrastructure support for this research was provided by the NIHR Imperial Biomedical Research Centre (BRC) to the NNRD.

Data were made available by the neoWONDER study supported by the National Institute for Health Research (NIHR) through an Advanced Fellowship awarded to CB (reference: NIHR300617).

This work was undertaken in the Office for National Statistics Secure Research Service using data from ONS and other owners and does not imply the endorsement of the ONS or other data owners.

We are grateful to the Department for Education for the provision of education data in the National Pupil Database.

