## Supplementary material for "Health and socioeconomic characteristics underlying educational attainment of children born preterm in England: a population cohort study using linked data"

Figure S4: Forest plot to show the unadjusted and adjusted associations between socio-economic and pre-term birth factors with educational outcomes (not meeting the expected level in KS1 writing or KS1 science) 21

Figure S5: Predicted probability of not meeting expected levels in educational outcomes across Index of Multiple Deprivation (IMD) at birth stratified by gestational age .... 22

References .... 23

**Description of the datasets**

The National Neonatal Research Database (NNRD) comprises a standard data extract (the Neonatal Data Set, an NHS Information Standard; DAPB1595) from Electronic Patient Records (EPR) very preterm infants. It contains data on all neonatal admissions from 2007 to the present, and approximately 450 de-identified entry items including demographic, diagnostic, daily, episodic and outcome variables.[1] All units in England have contributed data since 2012, providing whole population coverage for all preterm and sick infants admitted to neonatal units.[2]

The National Pupil Database (NPD) is a longitudinal administrative dataset collected and maintained by the Department for Education (DfE) of all children who attend state-funded schools in England.[3] This study used data on pupil characteristics, collected via the Pupil Level School Census three times per academic year (Autumn, Spring, and Summer terms), and educational attainment records. It includes the Early Years Foundation Stage Profile (EYFSP) at age 5 years, which is a summative assessment of a child's development that is used to support successful transition to year 1 of schooling and serves as a measure of school readiness. The NPD also includes a phonics check at age 6 years, and tests of reading, writing, mathematics, and science at age 7 years. All assessments are teacher-led.

Child-level linkage was performed by NHS Digital using personal identifiers (NHS numbers, date of birth, first name, surname, and postcodes) with subsequent deidentification, including removing name, date of birth, and exact date of neonatal discharge prior to study analysis.[4] Linkage to data from the Office for National Statistics provided information on registered deaths.

**Missing data and imputation**

Missingness was present for mode of delivery (8.5%), nutrition at discharge (6.5%), antenatal steroids (5.9%), and IMD (2.7%), Table S2. Prior to regression analyses, missing data on covariates were imputed using multiple imputation by chained equations, generating ten complete datasets under the assumption of missing at random. The imputation model incorporated all variables subsequently used in the regression analyses to preserve the multivariate relationships among covariates. Predictive mean matching or linear regression imputation was used for continuous variables, and logistic or multinomial regression for binary or categorial variables. Each imputed dataset was modelled independently, and results were pooled using Rubin's rule.

**Predictive performance of the fully adjusted models**

To evaluate out of sample predictive performance, the data were partitioned into training (70%) and test (30%) sets, stratifying by mother's ID to preserve clustering and prevent data leakage. The training set was multiply imputed (10 imputations), and GEE models were fitted on each complete dataset. Missing values in the test set were imputed using the median for continuous variables and the mode for categorical variables from the corresponding training set. Each trained model generated predictions on its paired test set and predicted probabilities for each participant were pooled by averaging across test sets. Performance was evaluated using AUC, sensitivity, specificity and accuracy.

**Extracted codes from NNRD to derive variables**

**1. Sepsis**

"An infant fulfilled criteria for sepsis if they had either a positive blood culture report of the growth of any organism from the National Neonatal Audit Programme (NNAP) late onset infection list of "Clearly pathogenic organisms" OR "other organisms" which includes coagulase negative Staphylococcus.[5] OR a discharge diagnosis indicating one or more "clearly" or "other" pathogenic organisms from the NNAP lists.

Discharge diagnoses that match NNAP list of "pure pathogens" AND "other organisms" in the NNRD field "Principal diagnosis at discharge" are:

sepsis - confirmed bacterial (gram positive)
sepsis / septicaemia - confirmed with +ve microbiology
e.coli sepsis / septicaemia
candida sepsis / septicaemia
group b streptococcal sepsis / septicaemia (gbs)
staphylococcal sepsis / septicaemia
staph. aureus sepsis / septicaemia
sepsis / septicaemia - specified - klebsiella sp.
sepsis / septicaemia - specified - enterobacter sp.
sepsis / septicaemia - specified - pseudomonas sp.
extended beta lactamase coliform infection/sepsis
listeria sepsis / septicaemia / disseminated
sepsis - confirmed bacterial (streptococci b positive)
sepsis - confirmed bacterial (streptococci positive)
streptococcal sepsis / septicaemia
salmonella sepsis
sepsis due to streptococcus
umbilical sepsis / septicaemia- group b streptococcus

**2. Infant of a diabetic mother**

To identify infants of mothers who had pre-existing or gestational diabetes:

Any of the following in ‘Principal diagnosis at discharge’:

“infant of diabetic mother"
"gestational diabetes” AND/OR

Any of the following in ‘Problems medical mother’:
“diabetes” AND/OR

Any of the following in ‘Problems during pregnancy:
“gestational diabetes”

**3. Hypertensive disorders of pregnancy (HDP)**

To identify infants of mothers with HDP, maternal HDP diagnoses were extracted from antenatal, delivery and neonatal NNRD data items.[6]

Any of the following in ‘Problems medical mother’:
“chronic hypertension” AND/OR

Any of the following in ‘Problems during pregnancy’:
“pregnancy-induced hypertension”
“pre-eclampsia”
“maternal HELLP” AND/OR

Any of the following in ‘Principal diagnosis at discharge’:
“mild pre-eclampsia”
“moderate pre-eclampsia”
“severe pre-eclampsia” AND/OR

Any of the following in ‘Drugs in labour’:
“antihypertensive”

**Table S1: Description of variables**

| Variable | Definition | Scale | Coding |
|---|---|---|---|
| ***School outcomes at age 5-7 years*** | | | |
| Early years Foundation Stage Profile (EYFSP) | Assessment at the end of Reception (age 5). Achieving at least the expected level in the Early Learning Goals related to communication and language, physical development, personal, social, and emotional development, literacy, and mathematics. | Binary | 0=achieved GLD, 1= non-GLD |
| Phonics | Assessment at the end of Year 1 (age 6). Pupils scoring >32 on the 40-word test are considered to have met the expected standard. Pupils who do not meet the expected standard retake the check again at the end of year 2. | Binary | 0= met the expected level, 1=did not meet the expected level |
| Key Stage 1 (KS1) | Assessment at the end of Year 2 (age 7) in reading, writing, mathematics, science. A child is assessed as meeting the expected level if they achieve level 2 or above. | Binary | 0= at/above expected level, 1= did not meet the expected level (per domain) |
| ***Child's characteristics*** | | | |
| Academic year of birth | Defined to align with the academic calendar year (1st September to 31st August inclusive the following year) to ensure that the cohort's attainment outcomes corresponded to the relevant academic year for the educational assessments. | Categorical | 0=2008/2009, 1=2009/2010, 2=2010/2011, 3=2011/2012 |
| Season of birth | Birth month is divided into three terms based on the academic year: Summer-born is defined as births during May-August inclusive; Spring-born as births during January-April; and Autumn-born as births during September-December | | 0=Summer-born; 1=Autumn-born; 2=Spring-born |
| Gestational age | Age at birth in completed weeks. | Continuous | 23-31 weeks inclusive |
| Gestational age group | Derived from the categorisation of the number of completed weeks of gestation at birth | Categorical | 0=27-31 weeks, 1=23-26 weeks |
| Birth-weight z-score | Weight z-scores were calculated using the LMS method of Cole and Green using sex, gestational age, postnatal age and birth weight.[16-18] | Continuous | |
| Multiple gestation | Derived from the number of fetuses at birth | Binary | 0=singleton, 1=multiple |
| Sex | Biological sex assigned at birth | Binary | 1=male, 0=female, |
| Small for gestational age (SGA) | <10th centile for birth weight | Binary | |
| Mode of delivery | Mother's mode of delivery | Binary | 0=C-section, 1=vaginal birth |
| ***Maternal characteristics*** | | | |
| Maternal age (years) | Age of mother at infant's birth | Continuous | |
| Smoking in pregnancy | Mother-confirmed smoking status during pregnancy | Binary | 0=not smoking, 1=smoking |

| | | | |
|---|---|---|---|
| Hypertensive disorders of pregnancy | See extracted codes in Supplement | Binary | |
| Infant of a diabetic mother | See extracted codes in Supplement | Binary | |
| ***Socio-economic/socio-demographic*** | ` | | |
| IMD decile | Index of Multiple Deprivation score: the decile measuring the deprivation score for the mother's location of residence at the time of the infant's birth. | Continuous | 1-10. 1 being the least deprived and 10 being the most deprived |
| IDACI decile | Income Deprivation Affecting Children Index: the decile measuring the income deprivation score for the child's location of residence in the year of entry to Reception. | Continuous | 1-10. 1 being the least deprived and 10 being the most deprived |
| Free school meal (FSM) eligibility | Child's FSM eligibility in the year of entry to Reception. | Binary | 0=not eligible, 1=eligible |
| Child's language | Child's major language group. Derived as the mode of entries across all Pupil Census records. | Binary | 0=English, 1=Not English |
| Maternal ethnicity | Ethnicities were self-reported and grouped according to the 2011 National Census categories[15] | Nominal | 0=White, 1=Asian, 2=Black, 3=Mixed, 4=Other |
| Child's ethnicity | Derived as the mode of entries across all Pupil Census records. | Nominal | 0=White, 1=Asian, 2=Black, 3=Chinese, 4=Mixed, 5=Other |
| ***Neonatal comorbidities*** | | | |
| Severe necrotising enterocolitis (NEC) | In accordance with the definition developed by Battersby et al.,[7] severe NEC is defined as NEC that required surgery or was confirmed at death. See extracted codes in Supplement. | Binary | 0=No severe NEC detected, 1 =severe NEC present |
| Treated retinopathy of prematurity (ROP) | Whether a baby was surgically treated or given drugs (Avastin) for ROP | Binary | 0 = No treatment given, 1 = treatment given |
| Sepsis | See extracted codes in Supplement | Binary | 0 =No sepsis detected, 1= Sepsis detected |
| Chronic Lung disease/bronchopulmonary dysplasia | Respiratory support at 36 weeks corrected gestational age | | 0 – No respiratory support given at 36 weeks, 1 – Respiratory support at 36 weeks |
| ***Neonatal brain injuries*** | | | |
| Cystic periventricular leukomalacia (CPVL) | CPVL on cerebral ultrasound scan (CUSS) &/or in final diagnosis record | Binary | 0=No CPVL of no record of CPVL, 1=CPVL detected |
| Porencephalic cyst | Porencephalic cyst visible on either the left or right hemispheres on CUSS &/or in final diagnosis record | Binary | 0=No porencephalic cyst of no record of porencephalic cyst, 1= porencephalic cyst detected |

| | | | |
|---|---|---|---|
| Hydrocephalus | Post-haemorrhagic hydrocephalus visible on CUSS &/or in final diagnosis record | Binary | 0=No hydrocephalus of no record of porencephalic cyst, 1= hydrocephalus detected |
| Intraventricular haemorrhage | The most severe grade of IVH seen on left/right hemisphere on the CUS scan &/or in final diagnosis record | Categorical | 0= no IVH, 1=IVH grade 1 or 2, 2=IVH grade 3 or 4 |
| ***Interventions*** | | | |
| Ante-natal corticosteroids | Whether a complete course of antenatal steroid courses given during pregnancy | Binary | 0=None or incomplete course, 1=complete course |
| Post-natal corticosteroids | Post-natal steroids received by 36 weeks' post-menstrual age | Binary | 0=No, 1=yes |
| ***Enteral nutrition at discharge from neo-natal unit*** | | | |
| Type of milk | Discharged on breast milk versus, formula, mixed, or other feeding. | Binary | 0=Breast milk, 1=Formula, 2=Mixed, 3=Other |

**Table S2: Distribution of missing data in covariates**

N=14,129

| Variable | Frequency | % missing |
|---|---|---|
| Vaginal birth | 1201 | 8.5 |
| Nutrition at discharge | 917 | 6.5 |
| Antenatal corticosteroids | 838 | 5.9 |
| IMD at birth | 386 | 2.7 |
| Postnatal corticosteroids | 305 | 2.2 |
| Maternal age | 45 | 0.3 |
| Major language | 15 | 0.1 |
| Birthweight z-score | <10 | <0.1 |
| Gestational age | 0 | 0 |
| Free school meal (5 years) | 0 | 0 |
| Necrotising enterocolitis | 0 | 0 |
| Bronchopulmonary dysplasia | 0 | 0 |
| Sepsis | 0 | 0 |
| Diabetic mother | 0 | 0 |
| Hypertensive disorder of pregnancy | 0 | 0 |
| Retinopathy of prematurity | 0 | 0 |
| Maternal smoking | 0 | 0 |
| Cystic periventricular leukomalacia | 0 | 0 |
| Intraventricular haemorrhage | 0 | 0 |
| Porencephalic cyst | 0 | 0 |
| Hydrocephalus | 0 | 0 |
| Sex | 0 | 0 |
| Multiple birth | 0 | 0 |
| Term of birth | 0 | 0 |

**Table S3: Characteristics of children with and without sufficient personal identifiers for linkage**

Frequencies (%) and median (IQR) are presented for categorical and continuous variables, respectively.

| | Insufficient personal identifiers for linkage | Identified for linkage | Total |
|---|---|---|---|
| N | **6,617 (27.7%)** | **17,245 (72.3%)** | **23,862 (100%)** |
| Mother's age at birth (years) | 30 (25 to 35) | 30 (25 to 35) | 30 (25 to 35) |
| Birth-weight z-score | -0.28 (-0.87 to 0.27) | -0.22 (-0.82 to 0.31) | -0.23 (-0.83 to 0.30) |
| Gestational age (completed weeks) | | | |
| 23 | 124 (1.9%) | 176 (1.0%) | 300 (1.3%) |
| 24 | 344 (5.2%) | 584 (3.4%) | 928 (3.9%) |
| 25 | 396 (6.0%) | 834 (4.8%) | 1,230 (5.2%) |
| 26 | 543 (8.2%) | 1,328 (7.7%) | 1,871 (7.8%) |
| 27 | 713 (10.8%) | 1,698 (9.8%) | 2,411 (10.1%) |
| 28 | 881 (13.3%) | 2,255 (13.1%) | 3,136 (13.1%) |
| 29 | 917 (13.9%) | 2,588 (15.0%) | 3,505 (14.7%) |
| 30 | 1,140 (17.2%) | 3,321 (19.3%) | 4,461 (18.7%) |
| 31 | 1,559 (23.6%) | 4,461 (25.9%) | 6,020 (25.2%) |
| Small for gestational age | 992 (15.0%) | 2,351 (13.6%) | 3,343 (14.0%) |
| Birth multiplicity | | | |
| Singleton | 4,729 (71.5%) | 12,547 (72.8%) | 17,276 (72.4%) |
| Multiple | 1,885 (28.5%) | 4,693 (27.2%) | 6,578 (27.6%) |
| Sex at birth | | | |
| Female | 3,067 (46.4%) | 8,010 (46.4%) | 11,077 (46.4%) |
| Male | 3,550 (53.6%) | 9,235 (53.6%) | 12,785 (53.6%) |
| IMD decile at birth | | | |
| 1 Most deprived | 974 (16.0%) | 3,025 (18.1%) | 3,999 (17.5%) |
| 2 | 808 (13.3%) | 2,462 (14.7%) | 3,270 (14.3%) |
| 3 | 819 (13.4%) | 2,101 (12.6%) | 2,920 (12.8%) |
| 4 | 711 (11.7%) | 1,739 (10.4%) | 2,450 (10.7%) |
| 5 | 568 (9.3%) | 1,616 (9.7%) | 2,184 (9.6%) |
| 6 | 503 (8.3%) | 1,360 (8.1%) | 1,863 (8.2%) |
| 7 | 458 (7.5%) | 1,256 (7.5%) | 1,714 (7.5%) |
| 8 | 436 (7.2%) | 1,152 (6.9%) | 1,588 (7.0%) |
| 9 | 414 (6.8%) | 1,030 (6.2%) | 1,444 (6.3%) |
| 10 Least deprived | 404 (6.6%) | 984 (5.9%) | 1,388 (6.1%) |
| Maternal ethnicity | | | |
| White | 4,280 (68.9%) | 12,462 (76.3%) | 16,742 (74.3%) |
| Asian | 868 (14.0%) | 1,806 (11.1%) | 2,674 (11.9%) |
| Black | 779 (12.5%) | 1,497 (9.2%) | 2,276 (10.1%) |
| Mixed | 118 (1.9%) | 263 (1.6%) | 381 (1.7%) |
| Other | 163 (2.6%) | 312 (1.9%) | 475 (2.1%) |
| Maternal smoking at birth | 1,129 (17.1%) | 3,067 (17.8%) | 4,196 (17.6%) |
| Complete course of ante-natal corticosteroids | 3,924 (66.8%) | 11,369 (70.1%) | 15,293 (69.2%) |
| Post-natal coticosteroids | 518 (8.1%) | 1,229 (7.3%) | 1,747 (7.5%) |
| Infant of a diabetic mother | 195 (2.9%) | 492 (2.9%) | 687 (2.9%) |
| Hypertensive disorders of pregnancy | 799 (12.1%) | 2,199 (12.8%) | 2,998 (12.6%) |

| | | | |
|---|---|---|---|
| Cystic periventricular leukomalacia | 95 (1.4%) | 277 (1.6%) | 372 (1.6%) |
| intraventricular haemorrhage (IVH) | | | |
| No IVH | 5,794 (87.6%) | 14,954 (86.7%) | 20,748 (86.9%) |
| IVH grade I-II | 567 (8.6%) | 1,788 (10.4%) | 2,355 (9.9%) |
| IVH grade III-IV | 256 (3.9%) | 503 (2.9%) | 759 (3.2%) |
| Porencephalic cyst | 57 (0.9%) | 229 (1.3%) | 286 (1.2%) |
| Hydrocephalus | 90 (1.4%) | 184 (1.1%) | 274 (1.1%) |
| Sepsis | 328 (5.0%) | 874 (5.1%) | 1,202 (5.0%) |
| Necrotising enterocolitis | 269 (4.1%) | 418 (2.4%) | 687 (2.9%) |
| Retinopathy of prematurity | 204 (3.1%) | 498 (2.9%) | 702 (2.9%) |
| Bronchopulmonary dysplasia | 1,864 (30.2%) | 4,636 (27.5%) | 6,500 (28.2%) |
| Enteral milk at discharge | | | |
| Breast | 1,697 (29.1%) | 4,776 (29.9%) | 6,473 (29.7%) |
| Formula | 2,173 (37.2%) | 6,213 (39.0%) | 8,386 (38.5%) |
| Other | 593 (10.2%) | 1,594 (10.0%) | 2,187 (10.0%) |
| Mixed | 1,376 (23.6%) | 3,364 (21.1%) | 4,740 (21.8%) |

**Table S4: Distribution (frequency/%) of children in the preterm cohort who did/did not meet expected levels in each outcome**

| | EYFSP (5 years) | | | Phonics (6 years) | | | KS1: Reading (7 years) | | | KS1: Writing (7 years) | | | KS1: Maths (7 years) | | | KS1: Science (7 years) | | |
|---|---|---|---|---|---|---|---|---|---|---|---|---|---|---|---|---|---|---|
| **Birth year** | **Met expected level** | **Did not meet expected level** | **TOTAL** | **Met expected level** | **Did not meet expected level** | **TOTAL** | **Met expected level** | **Did not meet expected level** | **TOTAL** | **Met expected level** | **Did not meet expected level** | **TOTAL** | **Met expected level** | **Did not meet expected level** | **TOTAL** | **Met expected level** | **Did not meet expected level** | **TOTAL** |
| **2008/2009** | 710 | 1,173 | 1,883 | 1,499 | 380 | 1,879 | 1,080 | 770 | 1,850 | 857 | 993 | 1,850 | 920 | 930 | 1,850 | 1,187 | 663 | 1,850 |
| | 37.7 | 62.3 | 100 | 79.8 | 20.2 | 100 | 58.4 | 41.6 | 100 | 46.3 | 53.7 | 100 | 49.7 | 50.3 | 100 | 64.2 | 35.8 | 100 |
| **2009/2010** | 1,615 | 2,247 | 3,862 | 3,032 | 802 | 3,834 | 2,199 | 1,600 | 3,799 | 1,798 | 2,001 | 3,799 | 1,942 | 1,857 | 3,799 | 2,427 | 1,372 | 3,799 |
| | 41.8 | 58.2 | 100 | 79.1 | 20.9 | 100 | 57.9 | 42.1 | 100 | 47.3 | 52.7 | 100 | 51.1 | 48.9 | 100 | 63.9 | 36.1 | 100 |
| **2010/2011** | 2,209 | 2,643 | 4,852 | 3,863 | 992 | 4,855 | 2,813 | 1,968 | 4,781 | 2,379 | 2,402 | 4,781 | 2,548 | 2,233 | 4,781 | 3,090 | 1,691 | 4,781 |
| | 45.5 | 54.5 | 100 | 79.6 | 20.4 | 100 | 58.8 | 41.2 | 100 | 49.8 | 50.2 | 100 | 53.3 | 46.7 | 100 | 64.6 | 35.4 | 100 |
| **2011/2012** | 2,392 | 2,539 | 4,931 | 3,884 | 1,052 | 4,936 | 2,695 | 2,016 | 4,711 | 2,318 | 2,393 | 4,711 | 2,514 | 2,196 | 4,710 | 2,988 | 1,723 | 4,711 |
| | 48.5 | 51.5 | 100 | 78.7 | 21.3 | 100 | 57.2 | 42.8 | 100 | 49.2 | 50.8 | 100 | 53.4 | 46.6 | 100 | 63.4 | 36.6 | 100 |
| **Total** | 6,926 | 8,602 | 15,528 | 12,278 | 3,226 | 15,504 | 8,787 | 6,354 | 15,141 | 7,352 | 7,789 | 15,141 | 7,924 | 7,216 | 15,140 | 9,692 | 5,449 | 15,141 |
| **4-year mean** | 43.4 | 56.6 | 100 | 79.3 | 20.7 | 100 | 58.1 | 41.9 | 100 | 48.2 | 51.8 | 100 | 51.9 | 48.1 | 100 | 64.0 | 36.0 | 100 |

**Table S5: Distribution of children in the preterm cohort who did not meet expected levels in each outcome by gestational age (birth years 2008/09 to 2011/12)**

| | EYFSP (5 years) | | Phonics (6 years) | | KS1 Reading (7 years) | | KS1 Writing (7 years) | | KS1 Maths (7 years) | | KS1 Science (7 years) | |
|---|---|---|---|---|---|---|---|---|---|---|---|---|
| Gestation (weeks) | n/N | % | n/N | % | n/N | % | n/N | % | n/N | % | n/N | % |
| 23 | 120/136 | 88.2 | 70/138 | 50.7 | 96/131 | 73.3 | 113/131 | 86.3 | 111/131 | 84.7 | 97/131 | 74 |
| 24 | 377/484 | 77.9 | 221/496 | 44.6 | 321/479 | 67 | 363/479 | 75.8 | 369/479 | 77 | 306/479 | 63.9 |
| 25 | 503/711 | 70.7 | 244/725 | 33.7 | 395/695 | 56.8 | 455/695 | 65.5 | 452/695 | 65 | 386/695 | 55.5 |
| 26 | 737/1170 | 63 | 296/1166 | 25.4 | 531/1132 | 46.9 | 657/1132 | 58 | 641/1132 | 56.6 | 491/1132 | 43.4 |
| 27 | 908/1506 | 60.3 | 335/1508 | 22.2 | 672/1463 | 45.9 | 804/1463 | 55 | 771/1463 | 52.7 | 595/1463 | 40.7 |
| 28 | 1206/2058 | 58.6 | 456/2047 | 22.3 | 864/1998 | 43.2 | 1063/1998 | 53.2 | 993/1998 | 49.7 | 761/1998 | 38.1 |
| 29 | 1242/2337 | 53.1 | 424/2333 | 18.2 | 912/2289 | 39.8 | 1142/2289 | 49.9 | 1038/2289 | 45.3 | 745/2289 | 32.5 |
| 30 | 1517/3043 | 49.9 | 542/3022 | 17.9 | 1091/2958 | 36.9 | 1364/2958 | 46.1 | 1240/2958 | 41.9 | 911/2958 | 30.8 |
| 31 | 1992/4083 | 48.8 | 638/4069 | 15.7 | 1472/3996 | 36.8 | 1828/3996 | 45.7 | 1601/3995 | 40.1 | 1157/3996 | 29 |

**Table S6: Unadjusted and adjusted GEE regression models for the association between socio-economic and preterm-birth factors with educational outcomes (KS1 writing and science) for children born 2008/09 to 2011/12 in England**

Results are expressed at odds ratios with 95% confidence intervals. The adjusted model includes all covariates listed for unadjusted models. N=14,129.

| | | KS1_Writing | | KS1_Science | |
|---|---|---|---|---|---|
| **Covariate** | **Freq.** | **Unadjusted** | **Adjusted** | **Unadjusted** | **Adjusted** |
| | | **OR/ 95% CI** | **OR/ 95% CI** | **OR/ 95% CI** | **OR/ 95% CI** |
| **Gestational age (completed weeks)** | | | | | |
| 23-24 | 544 | 3.99 | 2.59 | 4.53 | 2.62 |
| | | [3.23,4.93] | [2.01,3.34] | [3.74,5.50] | [2.07,3.33] |
| 25 | 634 | 2.18 | 1.56 | 2.93 | 1.9 |
| | | [1.82,2.60] | [1.27,1.91] | [2.46,3.49] | [1.55,2.34] |
| 26 | 1048 | 1.63 | 1.23 | 1.87 | 1.32 |
| | | [1.41,1.88] | [1.04,1.45] | [1.62,2.17] | [1.11,1.56] |
| 27 | 1363 | 1.44 | 1.14 | 1.68 | 1.26 |
| | | [1.26,1.64] | [0.99,1.31] | [1.47,1.93] | [1.08,1.46] |
| 28 | 1872 | 1.34 | 1.14 | 1.51 | 1.23 |
| | | [1.19,1.50] | [1.01,1.29] | [1.34,1.71] | [1.08,1.40] |
| 29 | 2158 | 1.21 | 1.13 | 1.21 | 1.09 |
| | | [1.08,1.35] | [1.00,1.27] | [1.08,1.37] | [0.96,1.24] |
| 30 | 2789 | 1.015 | 0.98 | 1.101 | 1.06 |
| | | [0.92,1.12] | [0.88,1.09] | [0.98,1.23] | [0.95,1.20] |
| 31: reference | 3721 | 1 | 1 | 1 | 1 |
| **Child characteristics** | | | | | |
| Birthweight z-score | 14,100 | 0.88 | 0.89 | 0.84 | 0.86 |
| | | [0.85,0.91] | [0.85,0.93] | [0.80,0.87] | [0.82,0.90] |
| Term of birth - Autumn: reference | 4382 | 1 | 1 | 1 | 1 |
| Term of birth - Spring | 4797 | 1.24 | 1.29 | 1.26 | 1.32 |
| | | [1.14,1.35] | [1.18,1.42] | [1.15,1.38] | [1.20,1.45] |
| Term of birth - Summer | 4950 | 1.69 | 1.84 | 1.68 | 1.83 |
| | | [1.55,1.84] | [1.68,2.01] | [1.54,1.84] | [1.67,2.01] |
| Singleton birth (vs. multiple birth) | 10,845 | 0.88 | 0.96 | 0.86 | 0.93 |
| | | [0.81,0.97] | [0.87,1.05] | [0.78,0.94] | [0.84,1.03] |
| Male (vs. female) | 7,493 | 1.69 | 1.83 | 1.24 | 1.31 |
| | | [1.58,1.81] | [1.70,1.96] | [1.16,1.33] | [1.21,1.41] |

| | | | | | |
|---|---|---|---|---|---|
| Vaginal birth (vs c-section) | 7,540 | 0.92 | 1.09 | 0.93 | 1.09 |
| | | [0.86,0.99] | [1.00,1.19] | [0.86,1.00] | [1.00,1.20] |
| Major language at home not English (vs. English) | 2,164 | 0.9 | 0.96 | 1.11 | 1.16 |
| | | [0.82,0.99] | [0.86,1.06] | [1.01,1.22] | [1.04,1.29] |
| **Socio-economic status** | | | | | |
| IMD quintile at birth 1 (most deprived) | 4,585 | 1.698 | 1.22 | 2.178 | 1.48 |
| | | [1.51,1.91] | [1.07,1.39] | [1.91,2.49] | [1.28,1.71] |
| IMD quintile 2 | 3,207 | 1.37 | 1.1 | 1.665 | 1.28 |
| | | [1.21,1.56] | [0.96,1.26] | [1.45,1.92] | [1.10,1.49] |
| IMD quintile 3 | 2,459 | 1.23 | 1.09 | 1.362 | 1.17 |
| | | [1.08,1.41] | [0.95,1.26] | [1.17,1.58] | [1.00,1.37] |
| IMD quintile 4 | 1,905 | 1.04 | 0.97 | 1.166 | 1.07 |
| | | [0.91,1.19] | [0.84,1.12] | [1.00,1.37] | [0.91,1.26] |
| IMD quintile at birth 5 (least deprived): reference | 1,587 | 1 | 1 | 1 | 1 |
| Free school meal eligible at 5 years | 3,354 | 2.03 | 1.65 | 2.2 | 1.78 |
| | | [1.87,2.20] | [1.50,1.80] | [2.02,2.38] | [1.62,1.95] |
| **Maternal characteristics** | | | | | |
| Maternal smoking at birth | 2,660 | 1.84 | 1.37 | 1.8 | 1.34 |
| | | [1.68,2.01] | [1.24,1.51] | [1.64,1.96] | [1.22,1.48] |
| Mother's age at birth (years) | 14,084 | 0.97 | 0.98 | 0.97 | 0.99 |
| | | [0.96,0.97] | [0.98,0.99] | [0.96,0.97] | [0.98,0.99] |
| Infant of a diabetic mother | 2,660 | 1.2 | 1.51 | 1.21 | 1.51 |
| | | [0.98,1.46] | [1.22,1.87] | [0.99,1.49] | [1.20,1.89] |
| Hypertensive disorder of pregnancy | 1,886 | 0.82 | 0.88 | 0.88 | 0.92 |
| | | [0.75,0.91] | [0.79,0.99] | [0.80,0.98] | [0.82,1.04] |
| **Interventions** | | | | | |
| Complete course of ante-natal corticosteroids | 9,285 | 0.87 | 0.88 | 0.86 | 0.86 |
| | | [0.80,0.93] | [0.82,0.96] | [0.79,0.93] | [0.79,0.94] |
| Post-natal corticosteroids | 937 | 2.42 | 1.26 | 2.57 | 1.21 |
| | | [2.10,2.79] | [1.06,1.50] | [2.25,2.94] | [1.03,1.43] |
| **Neonatal brain injuries** | | | | | |
| Cystic periventricular leukomalacia | 216 | 3.05 | 2.35 | 3.21 | 2.4 |
| | | [2.23,4.16] | [1.65,3.35] | [2.44,4.22] | [1.72,3.36] |
| No intra-ventricular haemorrhage (IVH): reference | 12,286 | 1 | 1 | 1 | 1 |
| IVH grade I-II | 1,473 | 1.26 | 0.99 | 1.38 | 1.05 |
| | | [1.13,1.41] | [0.88,1.12] | [1.24,1.54] | [0.93,1.19] |
| IVH grade III-IV | 370 | 2.88 | 1.48 | 3.14 | 1.53 |
| | | [2.29,3.62] | [1.13,1.94] | [2.55,3.86] | [1.17,1.99] |

| | | | | | |
|---|---|---|---|---|---|
| Porencephalic cysts | 179 | 3.06 | 1.69 | 2.94 | 1.43 |
| | | [2.18,4.31] | [1.15,2.49] | [2.18,3.97] | [0.99,2.06] |
| Hydrocephalus | 145 | 4.3 | 2.61 | 4.6 | 2.7 |
| | | [2.84,6.52] | [1.63,4.18] | [3.20,6.60] | [1.73,4.22] |
| **Neonatal morbidities** | | | | | |
| Sepsis | 688 | 1.48 | 1.09 | 1.7 | 1.18 |
| | | [1.26,1.72] | [0.93,1.29] | [1.46,1.98] | [1.00,1.39] |
| Necrotising enterocolitis | 302 | 2.74 | 1.84 | 3.06 | 1.96 |
| | | [2.12,3.55] | [1.40,2.41] | [2.42,3.86] | [1.53,2.52] |
| Retinopathy of prematurity | 383 | 3.17 | 1.5 | 3.31 | 1.42 |
| | | [2.49,4.02] | [1.15,1.95] | [2.67,4.09] | [1.11,1.82] |
| Bronchopulmonary dysplasia | 3,663 | 1.87 | 1.26 | 2.1 | 1.37 |
| | | [1.73,2.02] | [1.14,1.39] | [1.95,2.27] | [1.24,1.51] |
| **Nutrition at discharge** | | | | | |
| Breast milk: reference | 3,816 | 1 | 1 | 1 | 1 |
| Formula milk | 5,355 | 1.97 | 1.5 | 2.16 | 1.59 |
| | | [1.80,2.15] | [1.36,1.65] | [1.97,2.37] | [1.43,1.77] |
| Other milk | 1,334 | 2.4 | 1.62 | 2.75 | 1.71 |
| | | [2.11,2.73] | [1.41,1.87] | [2.42,3.13] | [1.48,1.97] |
| Mixed milk | 2,707 | 1.29 | 1.12 | 1.51 | 1.27 |
| | | [1.17,1.43] | [1.01,1.25] | [1.35,1.68] | [1.14,1.43] |
| **Out of sample predictive performance** | | | | | |
| AUC | | | 0.675 | | 0.685 |
| Sensitivity (positive class=failed to meet expected level) | | | 0.633 | | 0.631 |
| Specificity | | | 0.626 | | 0.645 |
| Accuracy | | | 0.629 | | 0.638 |
| Balanced accuracy | | | 0.630 | | 0.638 |

**Table S7: Sensitivity analysis substituting child's language for ethnicity: Fully adjusted GEE regression models for the association between socio-economic and preterm-birth factors with educational outcomes for children born 2008/09 to 2011/12 in England**

Model includes all other covariates reported in fully adjusted models in Table 2 and Table S4. N=14,129.

Results are expressed at odds ratios with 95% confidence intervals.

| | | EYFSP | Phonics | KS1_Reading | KS1_Writing | KS1_Maths | KS1_Science |
|---|---|---|---|---|---|---|---|
| | Freq. | OR/ 95% CI | OR/ 95% CI | OR/ 95% CI | OR/ 95% CI | OR/ 95% CI | OR/ 95% CI |
| **Child's ethnicity** | | | | | | | |
| White: reference | 10,421 | 1 | 1 | 1 | 1 | 1 | 1 |
| Asian | 1,336 | 1.24 | 0.92 | 0.98 | 0.87 | 0.88 | 1.15 |
| | | [1.08,1.41] | [0.78,1.09] | [0.86,1.11] | [0.77,0.99] | [0.78,1.01] | [1.00,1.31] |
| Black | 1,064 | 1 | 0.89 | 0.89 | 0.84 | 0.99 | 1.09 |
| | | [0.86,1.16] | [0.74,1.07] | [0.77,1.03] | [0.73,0.97] | [0.86,1.14] | [0.94,1.27] |
| Mixed | 984 | 0.81 | 0.8 | 0.88 | 0.92 | 0.98 | 0.93 |
| | | [0.70,0.94] | [0.67,0.96] | [0.75,1.02] | [0.79,1.06] | [0.84,1.13] | [0.80,1.09] |
| Other | 186 | 1.4 | 0.71 | 0.67 | 0.69 | 0.6 | 0.9 |
| | | [1.01,1.94] | [0.44,1.14] | [0.48,0.94] | [0.50,0.94] | [0.43,0.84] | [0.64,1.27] |

**Figure S1: Heatmap of correlations between outcomes and covariates**

Pearson's correlation coefficient was used to quantify the strength of the association between two continuous variables, the tetrachoric coefficient for two binary variables, and the point-biserial correlation coefficient for pairs of continuous and binary variables.

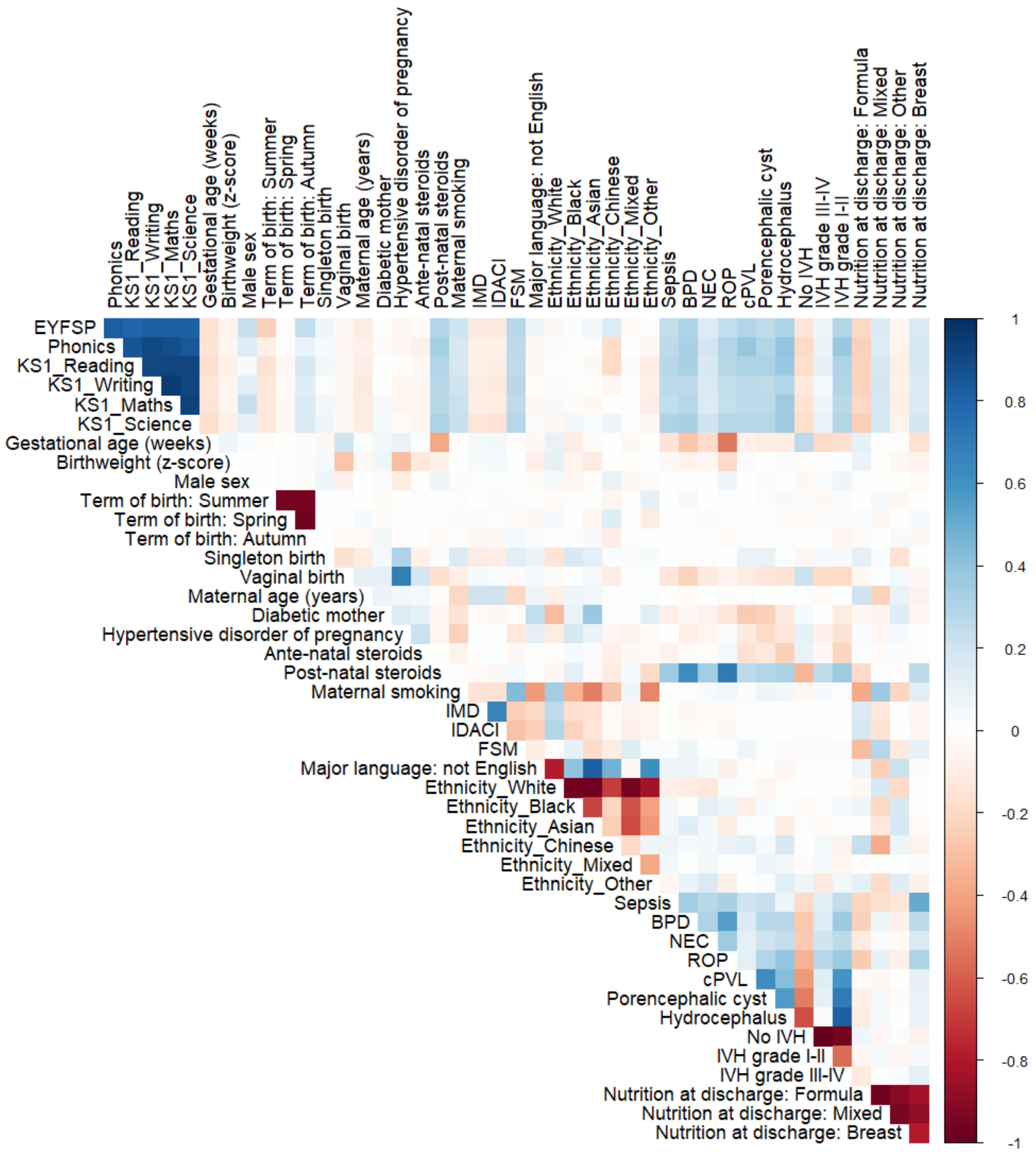

**Figure S2: Proportion of children who did not meet the expected level in outcomes by Income Deprivation affecting Children Index (IDACI) at age 5 years stratified by gestational age category (23-26 weeks and 27-31 weeks) and the overall cohort**

Trend tested using Cochran–Armitage test. P<0.001 for every line. IDACI 1=most deprived.

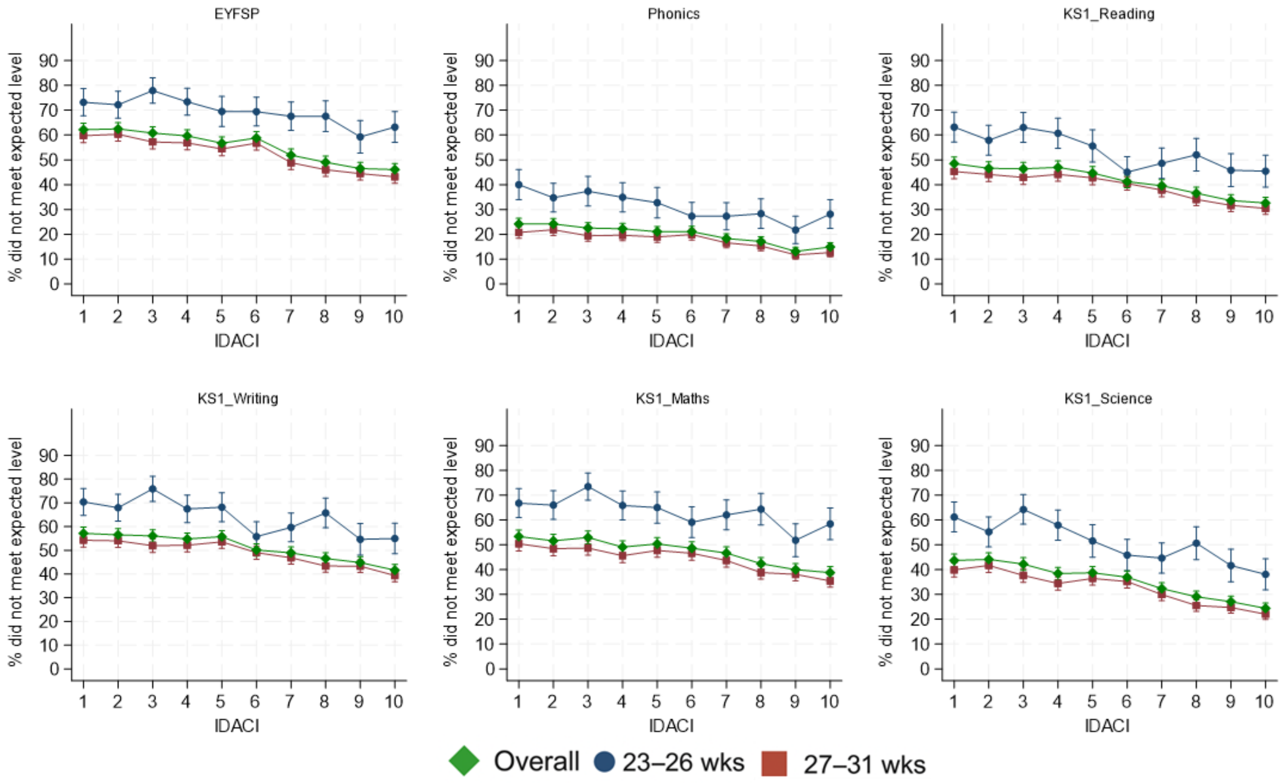

**Figure S3: Proportion of children who did not meet the expected level in outcomes by free school meal eligibility at age 5 years stratified by gestational age category (23-26 weeks and 27-31 weeks) and the overall cohort**

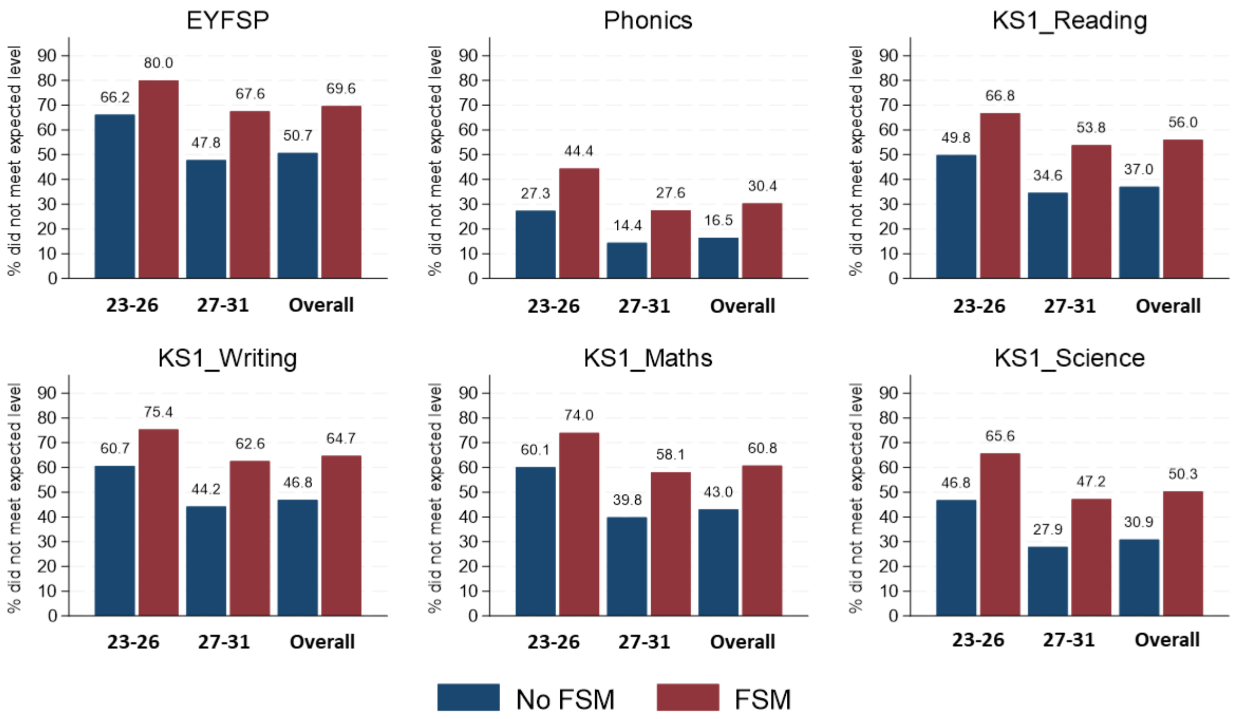

**Figure S4: Forest plot to show the unadjusted and adjusted associations between socio-economic and pre-term birth factors with educational outcomes (not meeting the expected level in KS1 writing or KS1 science)**

Odds ratio and 95% confidence intervals derived from GEE regression models.

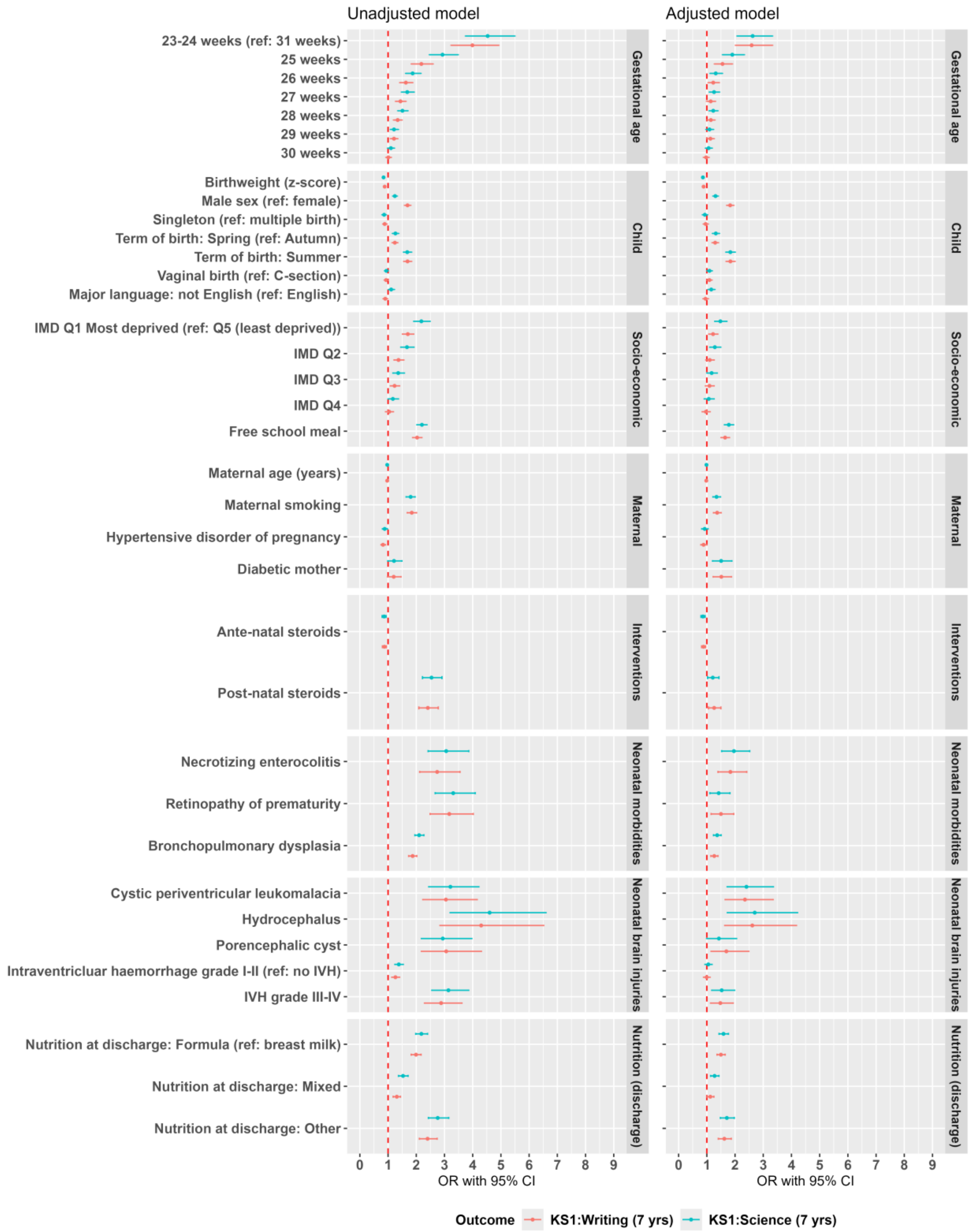

**Figure S5: Predicted probability of not meeting expected levels in educational outcomes across Index of Multiple Deprivation (IMD) at birth stratified by gestational age**

Predicted probabilities derived from GEE interaction model including gestational age (GA), IMD, and their interaction. The interaction term was insignificant in each model at a p-value of 5%. The parallel lines indicate that GA and deprivation additively increased the probability of poor attainment. IMD 1= most deprived.

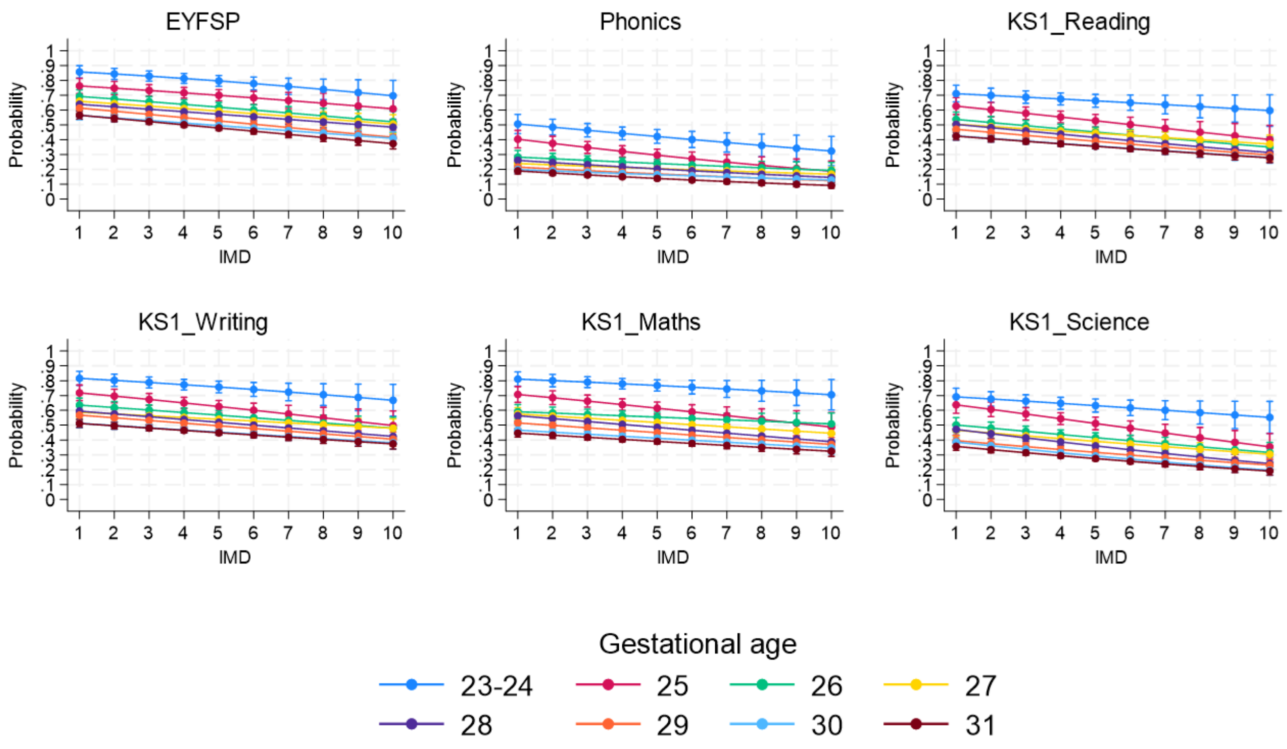
